# Change in Plasma Alpha-Tocopherol Associations with Attenuated Pulmonary Function Decline and with *CYP4F2* Missense Variation

**DOI:** 10.1101/2021.07.22.21260985

**Authors:** Jiayi Xu, Kristin A. Guertin, Nathan C. Gaddis, Anne H. Agler, Robert S. Parker, Jared M. Feldman, Alan R. Kristal, Kathryn B. Arnold, Phyllis J. Goodman, Catherine M. Tangen, Dana B. Hancock, Patricia A. Cassano

## Abstract

**Background:** Given its antioxidant activity, vitamin E is hypothesized to attenuate the age-related decline in pulmonary function.

**Objective:** We investigated the association between change in plasma vitamin E (ΔvitE) and pulmonary function decline and examined genetic and non-genetic factors associated with ΔvitE.

**Design:** We studied 1,144 men randomized to vitE in the Selenium and Vitamin E Cancer Prevention Trial. ΔvitE was calculated as the difference between baseline and year 3 vitE concentrations measured with gas chromatography-mass spectrometry. Pulmonary function (forced expiratory volume in the first second [FEV_1_]) was measured longitudinally with spirometry. We genotyped 555 participants (vitE-only arm) using the Illumina MEGA^ex^ array. We examined the association of ΔvitE with annual change in FEV_1_ using mixed-effects linear regression. We also examined the association of previously reported genetic and non-genetic factors with ΔvitE.

**Results:** Greater ΔvitE was associated with attenuated FEV_1_ decline, with stronger effects in adherent supplement responders: a 1 SD *higher* ΔvitE (+4 µmol/mmol free-cholesterol-adjusted α-tocopherol) attenuated FEV_1_ decline by ∼8.9 mL/year (*P*=0.014). This effect size is ∼1/4 of the effect of one year of aging, but in the opposite direction. The ΔvitE-FEV_1_ association was positive in never and current smokers (9.7 and 11.0 mL/year attenuated FEV_1_ decline, respectively), but there was little to no association in former smokers. *Greater* ΔvitE was associated with *lower* baseline α-tocopherol, *higher* baseline γ-tocopherol, *higher* baseline free cholesterol, European ancestry (vs. African ancestry) (all P<0.0001), and the minor allele of a missense variant in *CYP4F2* (rs2108622-T) (2.4 µmol/L *greater* ΔvitE; *P*=0.0032).

**Conclusions:** Greater response to vitE supplementation was associated with attenuated FEV_1_ decline, and this response was differed by rs2108622 such that individuals with the *C* allele may need a higher vitE intake dose to reach the same plasma level, compared to the *T* allele.

## Introduction

Vitamin E (vitE) is posited to have beneficial effects on lung health, given its role as a lipid-soluble antioxidant nutrient that scavenges free radicals in cells that can otherwise damage lung and other tissues (1, 2). Among the 8 forms of vitE, alpha-tocopherol (α-TOH) is the most abundant form in the circulation due to preferential post-absorption retention, while gamma-tocopherol (γ-TOH) is the major form in the U.S. diet (e.g., rich in vegetable oil and nuts). Positive association of vitE with cross-sectional pulmonary function (i.e., measured at a single time point) are reported in some (3-15), but not all (6, 11, 16-23) studies. Only two studies investigated associations with longitudinal decline in pulmonary function; a study of dietary intake of vitE with pulmonary function reported little to no association (4), while a study of serum vitE reported an association of lower baseline vitE with steeper decline in pulmonary function in heavy smokers (24). To our knowledge, the Respiratory Ancillary Study (RAS) to the Selenium and Vitamin E Cancer Prevention Trial (SELECT) is the only randomized controlled trial (RCT) to investigate the effect of vitE supplementation on pulmonary function decline. The RAS studied 2,920 men and, in an intention-to-treat analysis, reported a non-statistically significant (*P* = 0.19) protective effect of supplementation on decline (−33 mL/year in the vitE arm *versus* -39 mL/year in the placebo arm), but no evidence for an interaction of vitE supplementation with smoking status (25).

RCTs provide high-quality evidence to infer causal effects of nutrition on chronic disease outcomes, but intention-to-treat analyses have often failed to confirm nutrient-disease associations reported in observational studies and/or animal studies (26-28). While the RCT intention-to-treat design preserves randomization and a balance in participant characteristics by study arm (29), the intention-to-treat analysis may limit inferences for nutrition interventions if the intervention effect varies by individual characteristics including baseline nutriture and genetic variation. Thus, the efficacy of nutrient supplementation might vary by population subgroups, which would inform a precision nutrition approach, analogous to the concept of precision medicine (30).

An *in silico* simulation study reported that genes related to vitE metabolism are highly polymorphic (31) making it plausible that genetic differences contribute to inter-individual variation in response to supplementation. Genome-wide association studies (GWAS) have identified several genetic variants associated with cross-sectional plasma vitE concentration (32-34), and one study in men who were lifelong heavy smokers reported 2 genome-wide significant variants associated with plasma α-TOH concentration after 3 years of vitE supplementation (35).

We investigated the association of change in plasma vitE level (hereafter, ΔvitE), as a biomarker for the response to intervention with a supplement, with age-related decline in pulmonary function. We hypothesized that a greater response to vitE supplementation would lead to an attenuated pulmonary function decline in a generally healthy male population and tested this hypothesis in the RAS using a dose-response analysis. Furthermore, we hypothesized that genetic and non-genetic factors contribute to inter-individual differences in ΔvitE leading to sub-populations that benefit differentially from the same level of vitE supplementation.

## Methods

### Study Design and Participants

This study uses data from the RAS to SELECT, which was a phase 3 randomized controlled double-blinded trial of vitamin E and/or selenium supplementation for prostate cancer prevention. Details of the RAS and SELECT are published elsewhere (25, 36). Briefly, SELECT recruited 35,533 men aged ≥50 years for African Americans or > 55 years for all other men in the United States, Canada, and Puerto Rico. Eligibility criteria for SELECT included no prostate cancer diagnosis or suspicion of cancer, a serum prostate-specific antigen concentration ≤ 4 ng/mL, no more than 175 mg/day acetylsalicylic acid or 81 mg/day of acetylsalicylic acid if combined with clopidogrel bisulfate for anticoagulant therapy, a normal blood pressure, and no history of hemorrhagic stroke (36).

The RAS included men from 16 SELECT sites and preserved the randomized design. The four study arms were: vitE [400 IU/day of synthetic *all rac*-α-tocopheryl acetate] + selenium (Se) [200 μg/day of L-selenomethionine] (vitE+Se arm); vitE + Se placebo (vitE arm); vitE placebo + Se (Se arm); and vitE placebo + Se placebo (double placebo arm). The safety and efficacy of the supplementation have been considered in the original SELECT Trial (26). The primary endpoint of RAS was annual decline in FEV_1_; the secondary endpoint was annual decline in FEF25–75. These endpoints did not change significantly during the course of study across the intervention arms (25), which led us to the current exploratory analysis using the biomarker change as the exposure, instead of studying the effects of the intervention arms. The Cornell University IRB and the IRB at each of the 16 study sites approved the study. Analyses in this study focused on 1,144 participants of European or African ancestry in the two vitE (vitE or vitE+Se) arms with baseline (pre-intervention) and year 3 (on intervention) plasma vitE blood samples and at least 1 pulmonary function measurement (**Figure 1**).

**Figure 1.**
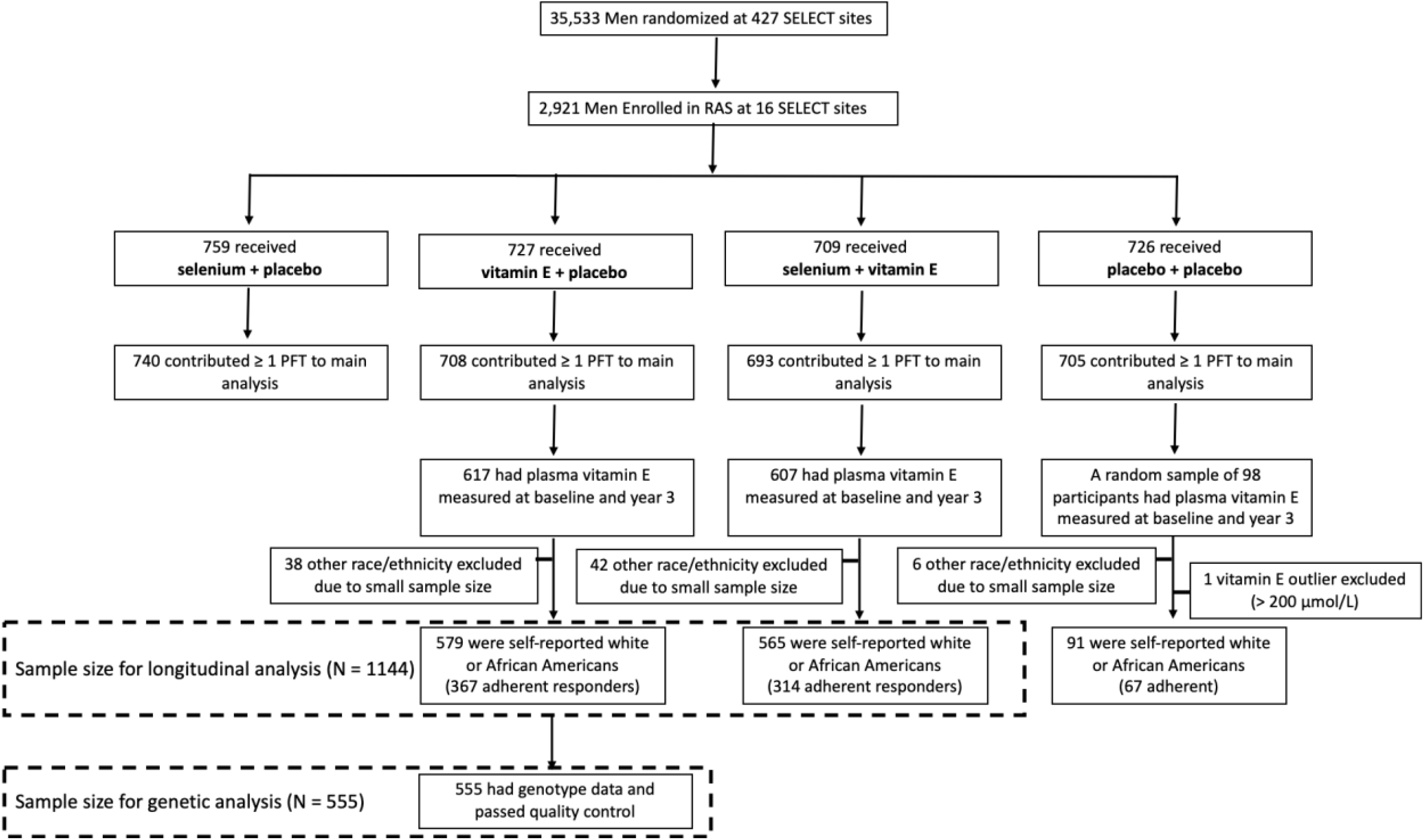
Flowchart of sample size included in this study

### Pulmonary Function

Pulmonary function, including forced expiratory volume in the first second (FEV_1_), was measured via the EasyOne handheld spirometer, which was previously tested for validity and reliability (37). The primary endpoint in the RAS was annual rate of change in FEV_1_. Pulmonary function tests (PFTs) were performed at specified SELECT visits in the RAS participants. Only PFTs meeting ATS standardization criteria (38) were included, and repeated PFTs had to be at least 2 years apart (from the first to the last PFT measure) to be included in our analyses.

### Vitamin E

Plasma vitE was assayed in samples from men in the vitE arms of the study and in a random sample of men in the double placebo arm. Blood samples were collected at SELECT baseline (prior to the start of supplements) and at the year 3 annual visit and stored at -80^°^C for a maximum of 9 years (see **Supplemental Methods**). Plasma α-TOH, γ-TOH and free un-esterified cholesterol were assayed via gas chromatography-mass spectrometry (GC-MS) on a Hewlett Packard 6890 gas chromatograph coupled with a Hewlett Packard 6890 mass spectrometer (Palo Alto, CA). Free cholesterol was measured as a proxy for total cholesterol given that the ratio of free to total cholesterol is relatively constant (39). Hereafter, free cholesterol-adjusted TOHs refer to the TOH levels after adjustment of free un-esterified cholesterol (i.e., TOH/cholesterol). Both raw TOH and free-cholesterol-adjusted TOH were used in this study because as a lipid-soluble vitamin, plasma vitE concentration varies by blood lipid levels (40).

### Genotyping

Genome-wide genotypes were assayed on a subset of unrelated RAS men in the vitE arm (*n* = 625). The vitE arm was selected for genotyping, rather than the vitE+Se arm, due to potential synergistic effects of Se supplementation. Participant samples of blood buffy coat, collected between 2004 and 2008 at the year 3 annual visit, were stored in a freezer at -80^°^C in the Human Metabolic Research Unit, Cornell University. In 2011, samples were shipped to the Center for Inherited Disease Research Laboratory, and DNA was extracted from peripheral blood leukocytes via the Puregene Blood Kit chemistry on an Autopure LS automated DNA purification instrument (Qiagen, Valencia, CA). In 2016, the Biotechnology Resource Center at Cornell University quantified the DNA concentrations using the Quantifluor dsDNA kit via the Spectra Max M2 plate reader (Molecular Devices, San Jose, CA), and samples were re-suspended in the hydration buffer (10mM Tris-Cl, 1mM EDTA, pH 8.0) to normalize concentrations, and then sent to the Northwest Genomics Center at University of Washington on dry ice in April 2016 for genotyping.

The Illumina Infinium^®^ Expanded Multi-Ethnic Genotyping Array (MEGA^ex^) (Illumina Inc., San Diego, CA, USA) was used to genotype more than 2 million genetic markers in 618 participants (7 participants were excluded due to very low DNA concentration). After excluding participants with failed call rate (*n* =15), genotype data were available for 440 participants of European ancestry [EAs] and 141 participants of African ancestry [AAs]. Quality control (QC) was applied on the genotyped individuals and the single nucleotide polymorphisms (SNPs). At the individual level, the QC filters included: (1) missing call rate > 3%, (2) duplicate sample (identity-by-state > 0.9), (3) first degree relative (identity-by-descent > 0.4), and (4) excessive homozygosity; there were 9 EAs and 1 AA excluded, leaving 431 EAs and 140 AAs. For the genetic analysis of plasma change in TOHs, the final sample size with all covariate information available was 555 (N_EA_ = 417, and N_AA_ = 138). At the SNP level, the QC filters included: (1) missing rate > 3%, (2) Hardy-Weinberg equilibrium p-value < 1×10^−4^ and (3) duplicate SNPs; these QC steps reduced the total genotyped SNP count from 2,036,060 to 1,671,215 for EAs and 1,662,290 for AAs. Genotype imputation was then performed based on the Haplotype Reference Consortium panel (version r1.1 2016) (41) using the Michigan Imputation Server. Given the sample size, we focused on variants with minor allele frequency (MAF) > 0.03 in both the ancestry-relevant 1000 Genomes Phase 3 super population (EUR for EAs and AFR for AAs) and the study population, resulting in a final set of 6,069,665 variants for EAs and 8,944,804 variants for AAs.

### Statistical Analysis of Plasma Vitamin E—Pulmonary Function Association

A dose-response analysis was used to test the association of change in plasma vitE biomarker levels (α-TOH and γ-TOH) in response to vitE supplementation (“ΔvitE” refers to change in α-TOH, the primary exposure of interest) with annual rate of change in FEV_1_ in the two vitE arms. A mixed-effects linear regression model was used to account for the repeated PFTs for each participant. The FEV_1_ value of each participant at each time point was the dependent variable in the model, with a variable for time as an independent variable that reflected the effect of one year of aging on change in pulmonary function (i.e., annual rate of change in FEV_1_). The association of ΔvitE with annual rate of change in FEV_1_ was modeled as an interaction term between ΔvitE and the time variable in the linear mixed-effects model (see equation below). The association was adjusted for covariates, including PFT-related factors such as age, height, ancestry, smoking status, cigarettes per day (current smokers only), as well as vitE status-related factors such as treatment arm (vitE arm, vitE+Se arm), baseline α-TOH, γ-TOH, and free cholesterol concentrations.

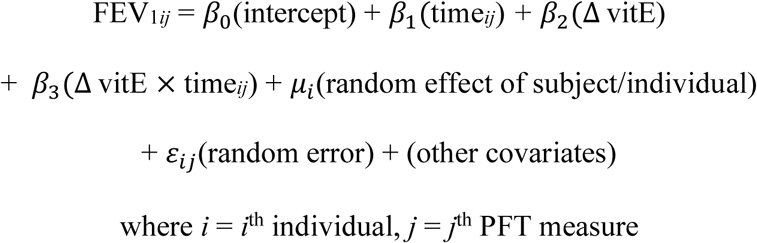

Sensitivity analyses explored subgroups where we hypothesized a stronger ΔvitE-FEV_1_ association. The first sensitivity analysis limited participants to those who were adherent to the intervention, defined as using ≥80% of the vitE supplement pills. Next, the analysis was further limited to adherent men who were responders to the vitE supplementation. Given that there was a decrease in plasma vitE concentration over 3 years, on average, in a subset of randomly selected participants in the placebo arm (*n* = 91; mean ΔvitE -3.8 µmol/L), responders to the vitE supplementation in the two intervention arms were defined as participants with a plasma ΔvitE value greater than the mean ΔvitE in the placebo arm. We chose the mean ΔvitE in the placebo arm as the reference point, instead of 0 (i.e., no change), because the change in plasma vitE over 3 years without a supplement is best estimated by the ΔvitE value in the placebo group (**Figure 2**). Results were robust to this choice.

**Figure 2.**
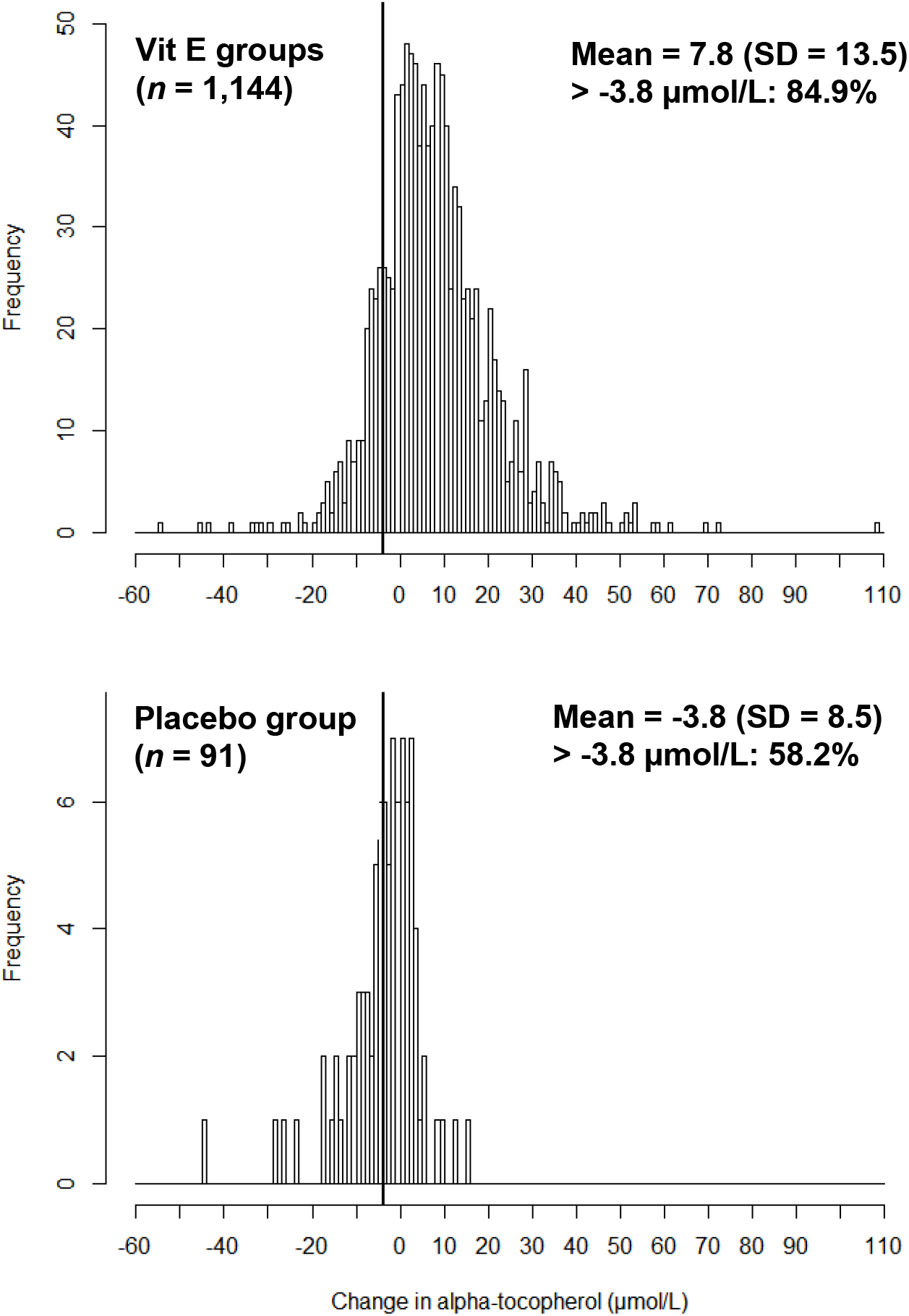
Frequency distribution of the change in plasma α-tocopherol (µmol/L) from baseline to year 3 in RAS. The black vertical line denotes the mean change in plasma α-tocopherol (−3.8 µmol/L) in the placebo arm. Vitamin E (vitE) groups include participants in the vitE supplementation only arm and vitE + Selenium supplementation arm. Responders in the vitE groups are defined as having a change in plasma α-tocopherol greater than -3.8 µmol/L. The mean and SD of change in plasma α-tocopherol is shown in each subfigure as well as the proportions of participants who had a change greater than -3.8 µmol/L.

The significance threshold for all analyses was set at two-sided *P* ≤ 0.05. Non-linearity was examined by including a square term for ΔvitE. The influence of one outlier for ΔvitE was examined by comparing the results with and without the outlier. Possible effect modification of the ΔvitE-FEV_1_ association by smoking status, by treatment arm, and by ancestry was examined in separate models via a 3-way interaction term (each putative effect modifier × ΔvitE × time).

### Statistical Analysis of Non-Genetic Factors on ΔvitE

We completed a systematic search of the literature for reports of non-genetic factors associated with plasma vitE concentration. For each factor identified, a further literature review sought to identify the biological plausibility of the putative association (**Supplemental methods**), leading to a final set of factors that were tested individually and then simultaneously in joint models. The variables tested were ancestry, intervention arm, and smoking status measured over the course of the study, and baseline values for age, BMI, α-TOH concentration, γ-TOH concentration, free cholesterol concentration, plasma Se concentration, and dietary intakes of vitamin C, fiber, alcohol. The dietary information was collected by a 120-item food frequency questionnaire (FFQ) administered at study baseline (42) to assess average food intake during the preceding 12 months, developed by the Nutrition Assessment Shared Resource (NASR) of Fred Hutchinson Cancer Research Center (Seattle, WA). Nutrient calculations were performed using the Nutrient Data System for Research (NDSR) software (version 2010), developed by the Nutrition Coordinating Center, University of Minnesota (Minneapolis, MN). For smoking status over the course of the study, we categorized the participants into never, former (stopped smoking prior to enrollment), intermittent (on and off smoking during the study), and persistent smokers to account for changes over the course of the study. We grouped intermittent and persistent smokers (i.e., current smokers during the study) to increase the power to detect associations. Linear regression models in SAS (version 9.4, SAS Institute, Cary, NC) tested the association of non-genetic factors with both raw ΔvitE and free-cholesterol-adjusted ΔvitE. The total R^2^ denoted the percent of variability in ΔvitE explained by the full model, and the individual R^2^ denoted the estimated variability explained by each individual factor (e.g., ancestry, baseline TOH concentration), which was calculated as the difference between total R^2^ with and without the factor in the model.

### Statistical Analysis of Genetic Variants on ΔvitE

We conducted agnostic (i.e., GWAS) analyses in order to identify SNPs associated with ΔvitE. We tested SNPs for associations under an additive model using imputed genotype dosage values (a fractional value between 0 and 2 indicating the expected number of minor allele copies) to account for any imputation uncertainty (43). GWAS analyses stratified by ancestry (N_EA_ = 417, N_AA_ = 138) were performed in rvtests (44) and then meta-analyzed using METAL (45) for each of four vitE phenotypes (raw ΔvitE, free-cholesterol-adjusted ΔvitE, raw Δγ-TOH, and free-cholesterol-adjusted Δγ-TOH) in models adjusted for age, baseline TOH concentration, and principal components to account for population substructure. We used the GWAS results to conduct *in-silico* look-up associations for two SNPs that were discovered in a previous GWAS of post-vitE-supplementation plasma α-TOH in heavy smokers (*P* < 5×10^−7^, accounting for 549,989 tested SNPs in the previous study) (35); we applied a Bonferroni-corrected p-value threshold of 0.025 (α=0.05/2 SNPs). Any SNPs that we replicated were further examined in models that included additional adjustment for the set of previously reported non-genetic covariates (**Supplemental Methods**) using SAS (version 9.4), which were not included in our parsimonious GWAS model that was designed to optimize statistical power.

## Results

### Participant Characteristics

1,144 participants were included in the model of annual rate of change in FEV_1_,: N_EA_ = 874 (76.4%) and N_AA_ = 270 (23.6%). The mean age of included participants was 63 years, and most participants had a history of smoking (49% former smokers, 19% current smokers) (**Table 1**) and were overweight (48%) or obese (34%).

**Table 1.**
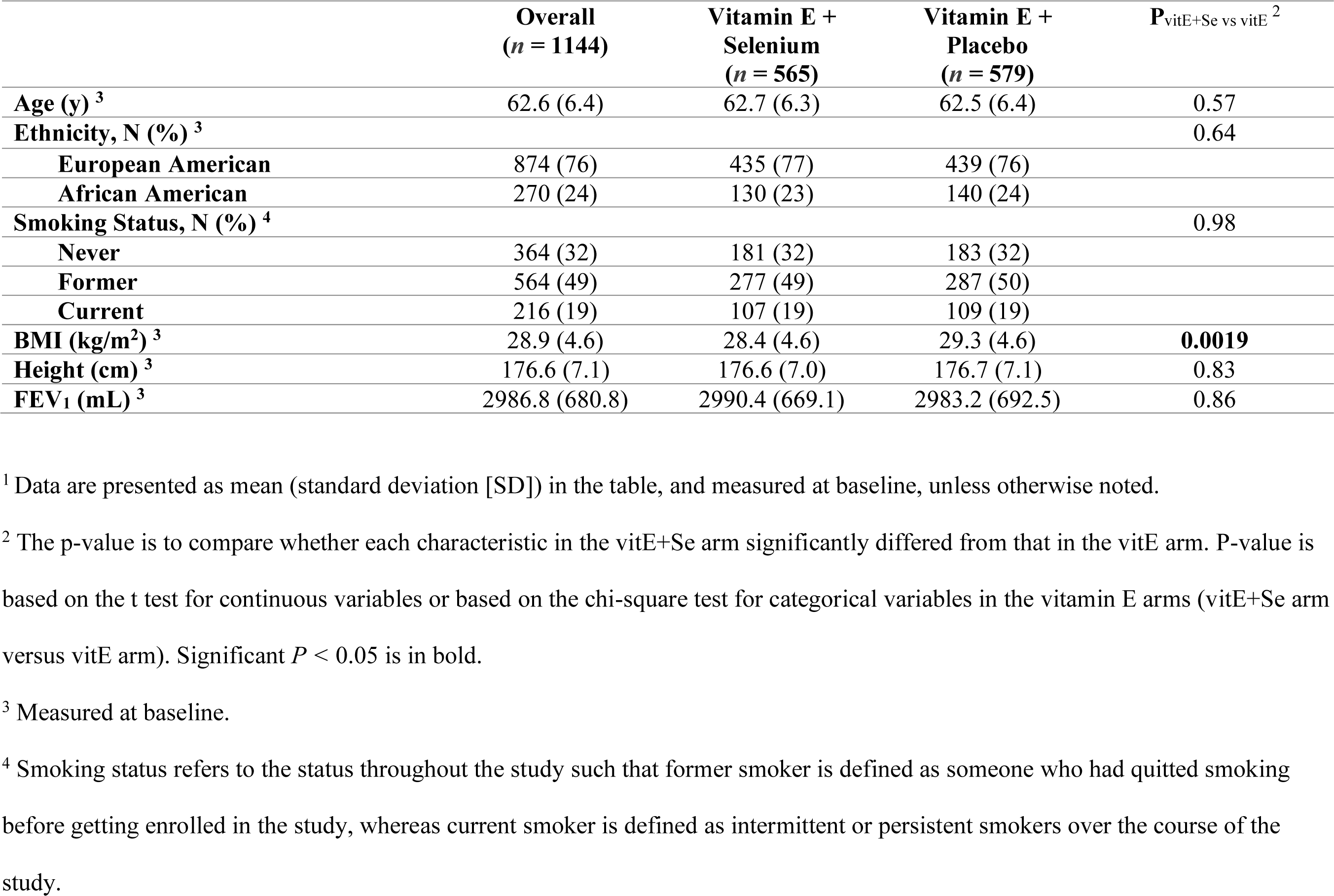
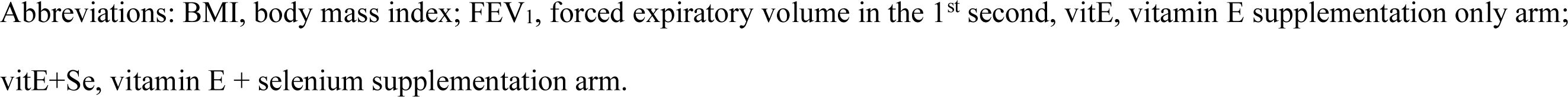
Characteristics of male participants with vitamin E supplementation in the Respiratory Ancillary Study (*n* = 1,144) ^1^.

|  | **Overall**<br>**($n = 1144$)** | **Vitamin E +**<br>**Selenium**<br>**($n = 565$)** | **Vitamin E +**<br>**Placebo**<br>**($n = 579$)** | **$P_{\text{vitE+Se vs vitE}}$<sup>2</sup>** |
| --- | --- | --- | --- | --- |
| **Age (y)**<sup>3</sup> | 62.6 (6.4) | 62.7 (6.3) | 62.5 (6.4) | 0.57 |
| **Ethnicity, N (%)**<sup>3</sup> |  |  |  | 0.64 |
| **European American** | 874 (76) | 435 (77) | 439 (76) |  |
| **African American** | 270 (24) | 130 (23) | 140 (24) |  |
| **Smoking Status, N (%)**<sup>4</sup> |  |  |  | 0.98 |
| **Never** | 364 (32) | 181 (32) | 183 (32) |  |
| **Former** | 564 (49) | 277 (49) | 287 (50) |  |
| **Current** | 216 (19) | 107 (19) | 109 (19) |  |
| **BMI (kg/m<sup>2</sup>)**<sup>3</sup> | 28.9 (4.6) | 28.4 (4.6) | 29.3 (4.6) | **0.0019** |
| **Height (cm)**<sup>3</sup> | 176.6 (7.1) | 176.6 (7.0) | 176.7 (7.1) | 0.83 |
| **FEV<sub>1</sub> (mL)**<sup>3</sup> | 2986.8 (680.8) | 2990.4 (669.1) | 2983.2 (692.5) | 0.86 |

<sup>1</sup> Data are presented as mean (standard deviation [SD]) in the table, and measured at baseline, unless otherwise noted.
<sup>2</sup> The p-value is to compare whether each characteristic in the vitE+Se arm significantly differed from that in the vitE arm. P-value is based on the t test for continuous variables or based on the chi-square test for categorical variables in the vitamin E arms (vitE+Se arm versus vitE arm). Significant $P < 0.05$ is in bold.
<sup>3</sup> Measured at baseline.
<sup>4</sup> Smoking status refers to the status throughout the study such that former smoker is defined as someone who had quit smoking before getting enrolled in the study, whereas current smoker is defined as intermittent or persistent smokers over the course of the study.
Abbreviations: BMI, body mass index; FEV<sub>1</sub>, forced expiratory volume in the 1<sup>st</sup> second, vitE, vitamin E supplementation only arm; vitE+Se, vitamin E + selenium supplementation arm.

On average, participants in the vitE arms showed an *increase* in plasma α-TOH after 3 years of vitE supplementation; the mean (SD) increase in plasma α-TOH was 8.2 (12.5) µmol/L in the vitE arm and 7.4 (14.5) µmol/L in the vitE+Se arm (**Supplemental Table 1**). In contrast, plasma α-TOH among participants in the double placebo arm (no vitE supplementation) decreased on average [mean Δ: -3.8 µmol/L α-TOH (SD = 8.5), *P* = 4.9×10^−5^, **Figure 2**].

Participants in the double placebo arm who reported use of pre-study vitE supplementation (>50 IU/d α-TOH or with other multivitamins) (17%) had a greater decrease in their plasma vitE level over 3 years (mean ΔvitE = -19 umol/L vs. -2.4 umol/L for participants not reporting pre-study vitE supplementation). For participants in the vitE arms, the increase in plasma vitE level was lower in participants who took pre-study vitE supplements (21%) (mean ΔvitE = +2.7 umol/L vs. +9.2 umol/L for those who did not take pre-study vitE supplementation).

Consistent with the change in plasma α-TOH, there was a notable change in plasma γ-TOH in participants in the vitE arms; plasma γ-TOH decreased by an average of 1.7 µmol/L in both vitE arms (SD = 2.4 and 2.6, respectively, in vitE arm and vitE+Se), but there was little to no change in the double placebo arm [mean Δ: +0.5 µmol/L (SD = 2.9), *P* = 0.10] (**Supplemental Table 1**). Six percent of participants were at risk of vitE deficiency at baseline using the clinical threshold of 12 µmol/L (46), and this dropped to 2% in the vitE arms after 3 years of supplementation. The two vitE intervention arms were similar in baseline characteristics and change in plasma tocopherol concentrations (**Table 1 and Supplemental Table 1**).

Although there was a statistically significant difference in BMI (*P* = 0.0019), the difference was not considered biologically meaningful with mean BMI of 29.3 in the vitE arm and 28.4 in the vitE+Se arm. While there was little to no correlation between baseline concentrations of α-TOH and γ-TOH in participants in the two vitE arms (Pearson *r* = 0.018, P=0.53), there was an inverse correlation between ΔvitE and Δγ-TOH (Pearson *r* = -0.22, P<0.0001).

The mean increase in ΔvitE after 3-years of supplementation was significantly higher in EAs compared with AAs in the vitE arm (PEA vs AA = 0.010, 9.0 µmol/L versus 5.9 µmol/L, respectively), with similar direction, but lower magnitude differences in the vitE+Se arm (PEA vs AA = 0.60, 7.5 µmol/L versus 6.9 µmol/L). The greater increase in plasma vitE levels in EAs was not related to differences by ancestry group in pre-study vitE supplement use (i.e., a greater increase due to less pre-study vitE use). Indeed, the proportion of EAs reporting pre-study vitE supplement use was greater (23%) compared to AAs (13%).

### Plasma ΔvitE and Longitudinal Pulmonary Function Decline

Plasma ΔvitE was associated with longitudinal FEV_1_ decline in adherent participants who responded to the vitE supplement (ΔvitE ≥-3.8 µmol/L) (**Table 2**), and there was little to no association of Δγ-TOH (**Supplemental Table 2**). In the full sample, an increase of 1 µmol/mmol in free-cholesterol-adjusted ΔvitE was associated with a 0.96 mL/year (SD=0.60) attenuation in annual FEV_1_ decline (*P* = 0.11). The effect size increased in sensitivity analyses as follows: men adherent to the intervention [increase of 1 µmol/mmol in free-cholesterol-adjusted ΔvitE associated with a +1.36 mL/year (SE = 0.78) attenuation in annual FEV_1_ decline, *P* = 0.082]; adherent participants who responded to the vitE supplementation [+2.22 mL/year (SD = 0.90), *P* = 0.014]. Overall, there was evidence that an increase in free-cholesterol-adjusted ΔvitE was associated with attenuation in FEV_1_ decline, with no evidence of non-linearity (*P* = 0.69). In adherent participants who responded to the intervention, a 1 SD greater ΔvitE (∼4 µmol/mmol free cholesterol) was associated with an attenuation of ∼9 mL/year in FEV_1_ decline. Consistent trends were observed for ΔvitE without adjusting free cholesterol (**Table 2**).

**Table 2.**
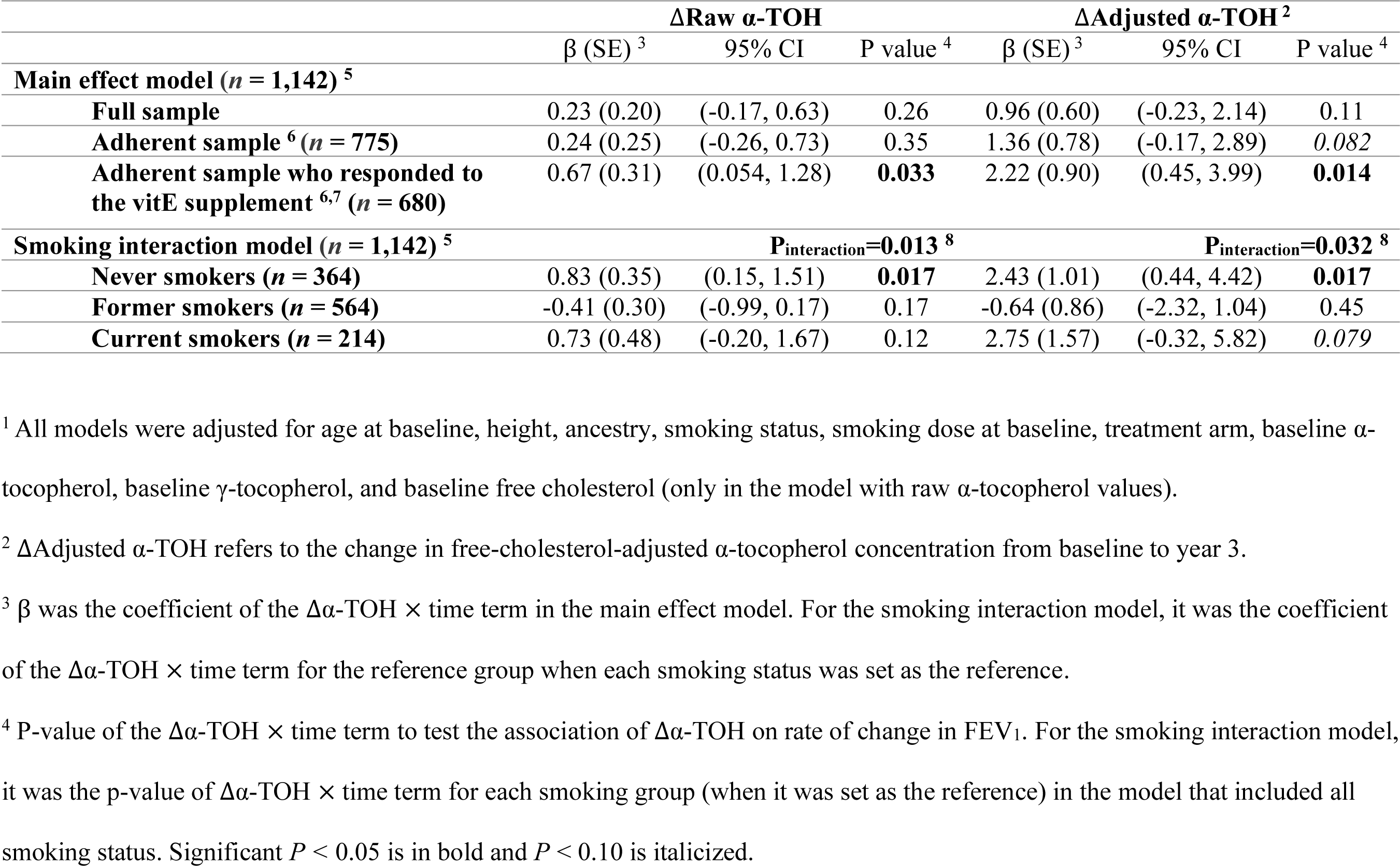
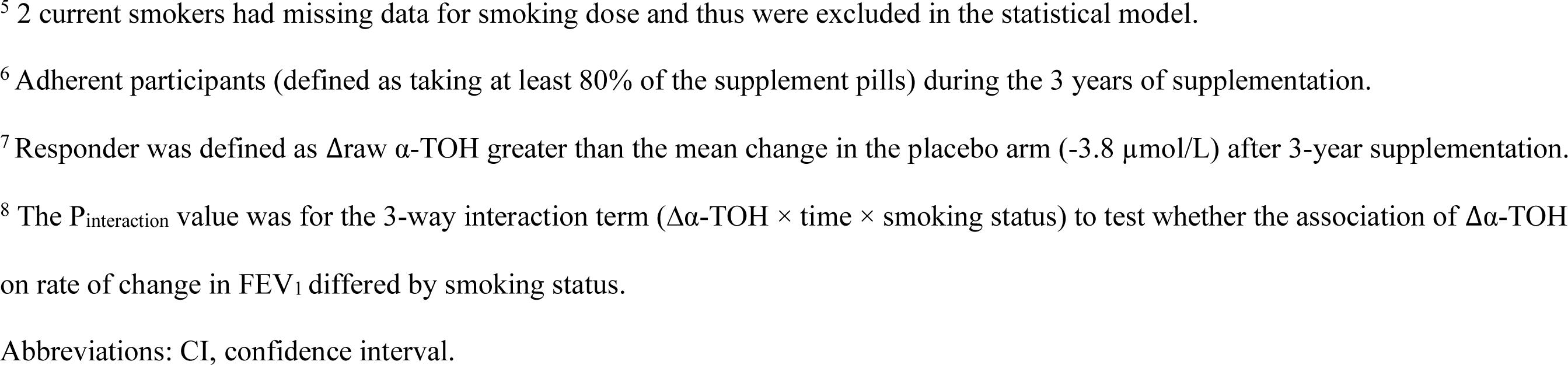
Association of plasma change in *α*-tocopherol after 3-year vitamin E supplementation, and its interaction with smoking status, on annual rate of change in FEV_1_ ^1^.

| | ΔRaw α-TOH | | | ΔAdjusted α-TOH<sup>2</sup> | | |
| --- | --- | --- | --- | --- | --- | --- |
| | $\beta$ (SE)<sup>3</sup> | 95% CI | P value<sup>4</sup> | $\beta$ (SE)<sup>3</sup> | 95% CI | P value<sup>4</sup> |
| **Main effect model ($n = 1,142$)<sup>5</sup>** |  |  |  |  |  |  |
| Full sample | 0.23 (0.20) | (-0.17, 0.63) | 0.26 | 0.96 (0.60) | (-0.23, 2.14) | 0.11 |
| Adherent sample<sup>6</sup> ($n = 775$) | 0.24 (0.25) | (-0.26, 0.73) | 0.35 | 1.36 (0.78) | (-0.17, 2.89) | *0.082* |
| Adherent sample who responded to the vitE supplement<sup>6,7</sup> ($n = 680$) | 0.67 (0.31) | (0.054, 1.28) | **0.033** | 2.22 (0.90) | (0.45, 3.99) | **0.014** |
| **Smoking interaction model ($n = 1,142$)<sup>5</sup>** |  |  |  |  |  |  |
|  | **P<sub>interaction</sub>=0.013<sup>8</sup>** |  |  | **P<sub>interaction</sub>=0.032<sup>8</sup>** |  |  |
| Never smokers ($n = 364$) | 0.83 (0.35) | (0.15, 1.51) | **0.017** | 2.43 (1.01) | (0.44, 4.42) | **0.017** |
| Former smokers ($n = 564$) | -0.41 (0.30) | (-0.99, 0.17) | 0.17 | -0.64 (0.86) | (-2.32, 1.04) | 0.45 |
| Current smokers ($n = 214$) | 0.73 (0.48) | (-0.20, 1.67) | 0.12 | 2.75 (1.57) | (-0.32, 5.82) | *0.079* |

<sup>1</sup> All models were adjusted for age at baseline, height, ancestry, smoking status, smoking dose at baseline, treatment arm, baseline α-tocopherol, baseline γ-tocopherol, and baseline free cholesterol (only in the model with raw α-tocopherol values).
<sup>2</sup> $\Delta$ Adjusted α-TOH refers to the change in free-cholesterol-adjusted α-tocopherol concentration from baseline to year 3.
<sup>3</sup> $\beta$ was the coefficient of the $\Delta\alpha$ -TOH × time term in the main effect model. For the smoking interaction model, it was the coefficient of the $\Delta\alpha$ -TOH × time term for the reference group when each smoking status was set as the reference.
<sup>4</sup> P-value of the $\Delta\alpha$ -TOH × time term to test the association of $\Delta\alpha$ -TOH on rate of change in FEV<sub>1</sub>. For the smoking interaction model, it was the p-value of $\Delta\alpha$ -TOH × time term for each smoking group (when it was set as the reference) in the model that included all smoking status. Significant $P < 0.05$ is in bold and $P < 0.10$ is italicized.
<sup>5</sup> 2 current smokers had missing data for smoking dose and thus were excluded in the statistical model.
<sup>6</sup> Adherent participants (defined as taking at least 80% of the supplement pills) during the 3 years of supplementation.
<sup>7</sup> Responder was defined as $\Delta$ raw α-TOH greater than the mean change in the placebo arm ( $-3.8 \mu\text{mol/L}$ ) after 3-year supplementation.
<sup>8</sup> The $P_{\text{interaction}}$ value was for the 3-way interaction term ( $\Delta\alpha\text{-TOH} \times \text{time} \times \text{smoking status}$ ) to test whether the association of $\Delta\alpha\text{-TOH}$ on rate of change in $\text{FEV}_1$ differed by smoking status.
Abbreviations: CI, confidence interval.

Smoking status modified the association of plasma ΔvitE with FEV_1_ decline (Psmoking interaction < 0.05, **Table 2)**. An increase in free-cholesterol-adjusted ΔvitE was significantly associated with attenuation in FEV_1_ decline in never smokers (*P* = 0.017) with a similar but not statistically significant trend in current smokers (*P* = 0.079), and, contrary to expectation, little to no association in former smokers (*P* = 0.45, **Table 2**). To put the effect magnitude in context, 1 SD higher free-cholesterol-adjusted ΔvitE (∼4 µmol/mmol free cholesterol) was associated with 9.7 mL/yr and 11.0 mL/yr attenuation in FEV_1_ decline in never and current smokers, respectively. The same trend was observed for raw ΔvitE (**Table 2**), and in sensitivity analysis limited to adherent men (**Supplemental Table 3**). Neither ancestry (AA, EA) nor treatment arm (vitE, vitE+Se) modified the association of ΔvitE with FEV_1_ decline (P_interaction_ = 0.85 and 0.43 for ancestry and treatment arm, respectively).

**Table 3.**
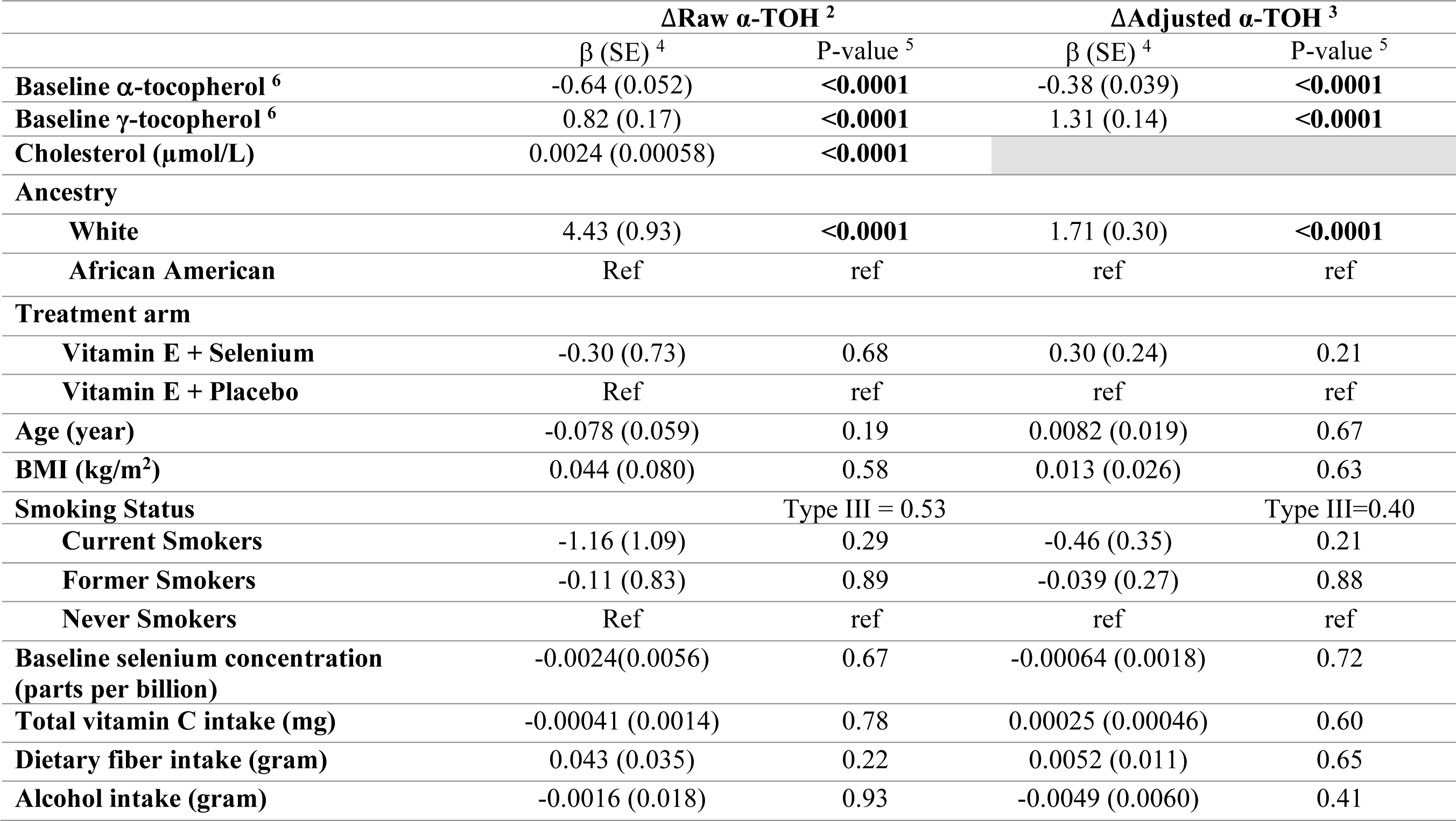
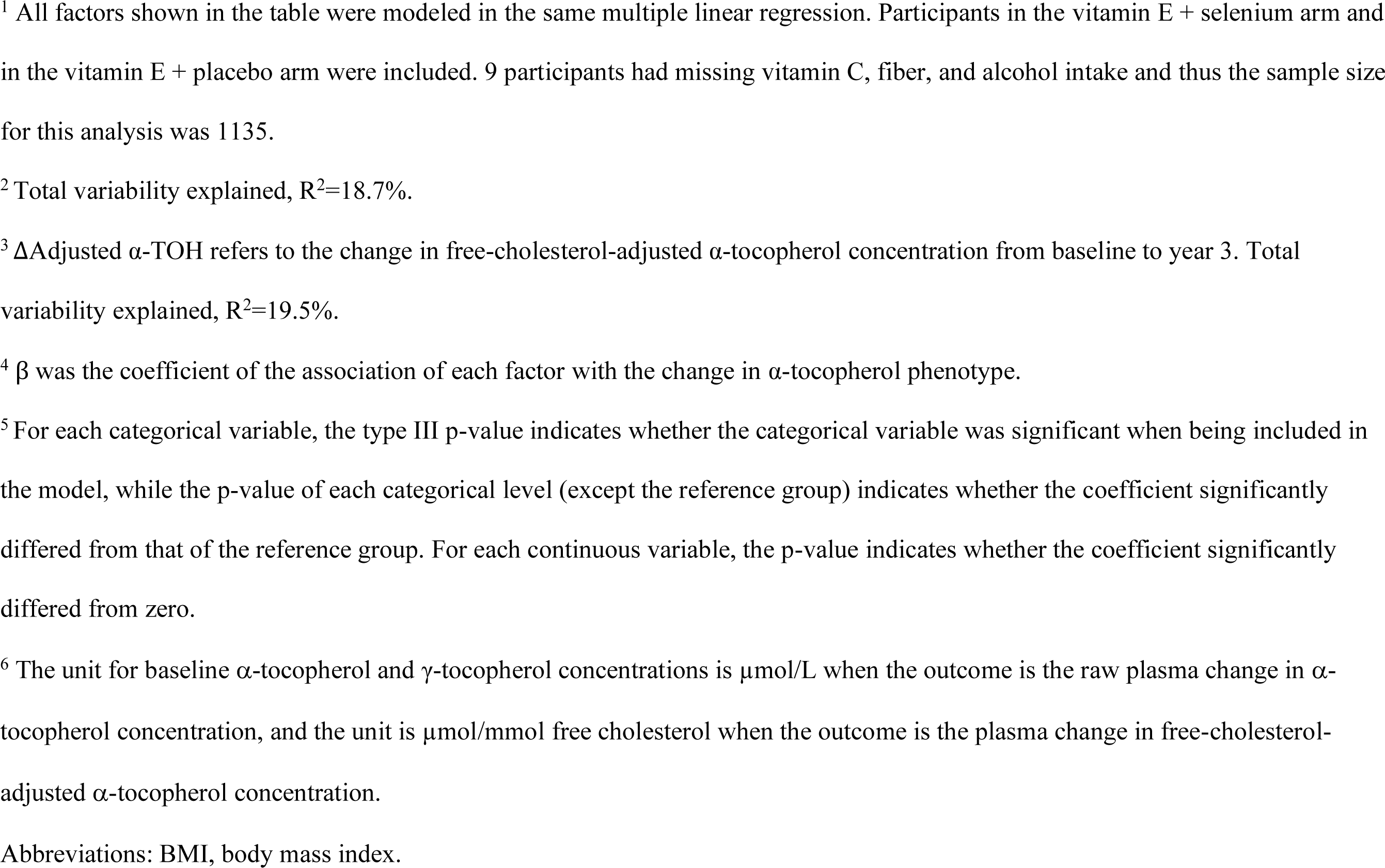
Factors associated with plasma change in *α*-tocopherol after 3-year vitamin E supplementation in the Respiratory Ancillary Study (*n* = 1,135) ^1^.

| | ΔRaw α-TOH <sup>2</sup> | | ΔAdjusted α-TOH <sup>3</sup> | |
| --- | --- | --- | --- | --- |
| | $\beta$ (SE) <sup>4</sup> | P-value <sup>5</sup> | $\beta$ (SE) <sup>4</sup> | P-value <sup>5</sup> |
| **Baseline α-tocopherol <sup>6</sup>** | -0.64 (0.052) | **<0.0001** | -0.38 (0.039) | **<0.0001** |
| **Baseline γ-tocopherol <sup>6</sup>** | 0.82 (0.17) | **<0.0001** | 1.31 (0.14) | **<0.0001** |
| **Cholesterol (μmol/L)** | 0.0024 (0.00058) | **<0.0001** |  |  |
| **Ancestry** |  |  |  |  |
| **White** | 4.43 (0.93) | **<0.0001** | 1.71 (0.30) | **<0.0001** |
| **African American** | Ref | ref | ref | ref |
| **Treatment arm** |  |  |  |  |
| **Vitamin E + Selenium** | -0.30 (0.73) | 0.68 | 0.30 (0.24) | 0.21 |
| **Vitamin E + Placebo** | Ref | ref | ref | ref |
| **Age (year)** | -0.078 (0.059) | 0.19 | 0.0082 (0.019) | 0.67 |
| **BMI ($\text{kg}/\text{m}^2$)** | 0.044 (0.080) | 0.58 | 0.013 (0.026) | 0.63 |
| **Smoking Status** |  | Type III = 0.53 |  | Type III=0.40 |
| **Current Smokers** | -1.16 (1.09) | 0.29 | -0.46 (0.35) | 0.21 |
| **Former Smokers** | -0.11 (0.83) | 0.89 | -0.039 (0.27) | 0.88 |
| **Never Smokers** | Ref | ref | ref | ref |
| **Baseline selenium concentration (parts per billion)** | -0.0024(0.0056) | 0.67 | -0.00064 (0.0018) | 0.72 |
| **Total vitamin C intake (mg)** | -0.00041 (0.0014) | 0.78 | 0.00025 (0.00046) | 0.60 |
| **Dietary fiber intake (gram)** | 0.043 (0.035) | 0.22 | 0.0052 (0.011) | 0.65 |
| **Alcohol intake (gram)** | -0.0016 (0.018) | 0.93 | -0.0049 (0.0060) | 0.41 |

<sup>1</sup> All factors shown in the table were modeled in the same multiple linear regression. Participants in the vitamin E + selenium arm and in the vitamin E + placebo arm were included. 9 participants had missing vitamin C, fiber, and alcohol intake and thus the sample size for this analysis was 1135.
<sup>2</sup> Total variability explained, $R^2=18.7\%$ .
<sup>3</sup> $\Delta$ Adjusted α-TOH refers to the change in free-cholesterol-adjusted α-tocopherol concentration from baseline to year 3. Total variability explained, $R^2=19.5\%$ .
<sup>4</sup> $\beta$ was the coefficient of the association of each factor with the change in α-tocopherol phenotype.
<sup>5</sup> For each categorical variable, the type III p-value indicates whether the categorical variable was significant when being included in the model, while the p-value of each categorical level (except the reference group) indicates whether the coefficient significantly differed from that of the reference group. For each continuous variable, the p-value indicates whether the coefficient significantly differed from zero.
<sup>6</sup> The unit for baseline α-tocopherol and γ-tocopherol concentrations is $\mu\text{mol/L}$ when the outcome is the raw plasma change in α-tocopherol concentration, and the unit is $\mu\text{mol}/\text{mmol}$ free cholesterol when the outcome is the plasma change in free-cholesterol-adjusted α-tocopherol concentration.
Abbreviations: BMI, body mass index.

### Non-Genetic Factors and Plasma ΔvitE

Plasma ΔvitE values ranged from -55 µmol/L to +109 µmol/L in the two vitE arms [**Figure 2**, mean (SD) of 7.8 (13.5), with no difference between arms, *P* = 0.68]. *Higher* α-TOH at the study baseline was associated with a *lower* increase in ΔvitE (*P* < 0.0001), while *higher* free cholesterol and *higher* γ-TOH at baseline were associated with a *greater* increase in ΔvitE (*P* < 0.0001) (**Table 3**). Ancestry was associated with ΔvitE (*P* < 0.0001), with a smaller increase in ΔvitE in African American men and generally this pattern persisted regardless of smoking status (**Supplemental Table 4**). There was little to no association between age, BMI, smoking status, and other nutrition factors, including baseline plasma Se concentration, dietary intake of vitamin C, fiber, and alcohol in relation to ΔvitE (*P* > 0.05). Overall, we saw consistent findings between raw and free-cholesterol-adjusted Δα-TOH (**Table 3**). The fully adjusted model explained 18.7% of the variability (R^2^) in ΔvitE in the two vitamin E arms (*n* = 1,144), and baseline concentrations of α-TOH, γ-TOH, free cholesterol and ancestry explained 10.9%, 1.7%, 1.2%, and 1.6% of the variability, respectively.

**Table 4.**
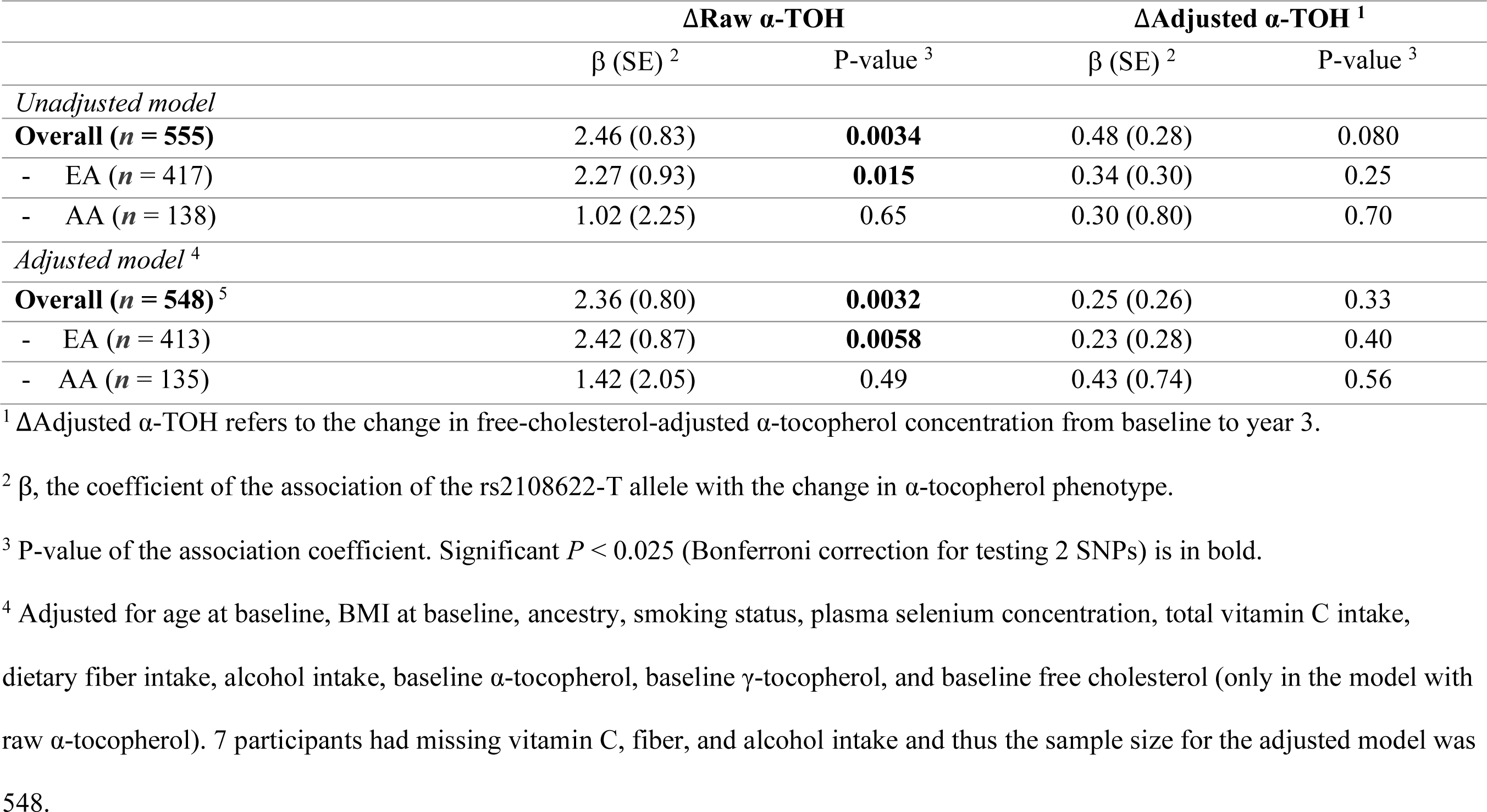
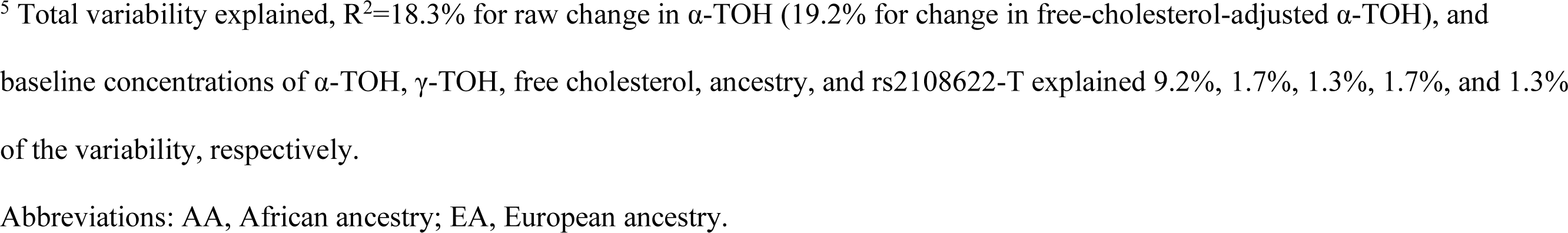
The association of rs2108622-T with plasma change in *α*-tocopherol for male participants randomized to the vitamin E only arm (400 IU/day *all rac-α*-tocopherol) in the Respiratory Ancillary Study (*n* = 555)

| | ΔRaw α-TOH | | ΔAdjusted α-TOH <sup>1</sup> | |
| --- | --- | --- | --- | --- |
| | $\beta$ (SE) <sup>2</sup> | P-value <sup>3</sup> | $\beta$ (SE) <sup>2</sup> | P-value <sup>3</sup> |
| *Unadjusted model* |  |  |  |  |
| **Overall ($n = 555$)** | 2.46 (0.83) | **0.0034** | 0.48 (0.28) | 0.080 |
| - EA ($n = 417$) | 2.27 (0.93) | **0.015** | 0.34 (0.30) | 0.25 |
| - AA ($n = 138$) | 1.02 (2.25) | 0.65 | 0.30 (0.80) | 0.70 |
| *Adjusted model <sup>4</sup>* |  |  |  |  |
| **Overall ($n = 548$) <sup>5</sup>** | 2.36 (0.80) | **0.0032** | 0.25 (0.26) | 0.33 |
| - EA ($n = 413$) | 2.42 (0.87) | **0.0058** | 0.23 (0.28) | 0.40 |
| - AA ($n = 135$) | 1.42 (2.05) | 0.49 | 0.43 (0.74) | 0.56 |

<sup>1</sup> $\Delta$ Adjusted α-TOH refers to the change in free-cholesterol-adjusted α-tocopherol concentration from baseline to year 3.
<sup>2</sup> $\beta$ , the coefficient of the association of the rs2108622-T allele with the change in α-tocopherol phenotype.
<sup>3</sup> P-value of the association coefficient. Significant $P < 0.025$ (Bonferroni correction for testing 2 SNPs) is in bold.
<sup>4</sup> Adjusted for age at baseline, BMI at baseline, ancestry, smoking status, plasma selenium concentration, total vitamin C intake, dietary fiber intake, alcohol intake, baseline α-tocopherol, baseline γ-tocopherol, and baseline free cholesterol (only in the model with raw α-tocopherol). 7 participants had missing vitamin C, fiber, and alcohol intake and thus the sample size for the adjusted model was 548.
<sup>5</sup> Total variability explained, $R^2=18.3\%$ for raw change in α-TOH (19.2% for change in free-cholesterol-adjusted α-TOH), and baseline concentrations of α-TOH, γ-TOH, free cholesterol, ancestry, and rs2108622-T explained 9.2%, 1.7%, 1.3%, 1.7%, and 1.3% of the variability, respectively.
Abbreviations: AA, African ancestry; EA, European ancestry.

### Genetic Factors and Plasma ΔvitE

GWAS analyses revealed no genome-wide significant loci (P < 5×10^−8^) for change in TOH concentration (either α- or γ-TOH) in the vitE arm (results not shown). We used the GWAS results to study 2 SNPs (rs964184 and rs2108622) that were previously reported in a GWAS of circulating α-TOH measured after 3-years of vitE supplementation in male current smokers, but were not tested for replication (35). We replicated one SNP: rs2108622 was associated with ΔvitE after 3 years of vitE supplementation in the healthy male participants in the RAS, with *P*-value passing the Bonferroni-corrected threshold for replication (P < 0.025). Rs2108622, a missense SNP in the cytochrome P450 family 4 subfamily F member 2 (*CYP4F2*) gene on chromosome 19, was directly genotyped on the MEGA^ex^ array, so its dosages were 0 (major allele homozygote), 1 (heterozygote), and 2 (minor allele homozygote). When the genotype was modeled as a categorical variable, heterozygotes for the rs2108622 minor allele (*CT* genotype) had a +1.75 μmol/L ΔvitE, compared to homozygotes for the major allele (*CC* genotype), whereas homozygotes for the minor allele (*TT* genotype) had a +5.66 μmol/L ΔvitE, compared to *CC* genotype.

The mean ΔvitE for participants with 0, 1, 2 minor alleles of rs2108622 was 7.6 µmol/L, 9.5 µmol/L, 12.7 µmol/L in EAs, and ancestry-specific analyses revealed a consistent direction of association for rs2108622 (P = 0.61 for interaction with ancestry; mean ΔvitE for 0, 1, 2 minor alleles was 5.8 µmol/L, 6.0 µmol/L, 12.9 µmol/L in AAs). Thus, findings with the two ancestries combined are presented herein. In the fully adjusted model where the number of SNP alleles was modelled as a continuous variable, each additional copy of the rs2108622-*T* minor allele (frequency = 31% in EAs and 13% in AAs) was associated with 2.36 µmol/L *greater* increase in ΔvitE (P = 0.0032, **Table 4**), compared to the *C* major allele. This effect magnitude is ∼20% of 1 SD of ΔvitE in the vitE arm (SD = 12.5 µmol/L). In addition, this SNP explained about a third of the variability (R^2^) in ΔvitE that was initially explained by ancestry (without rs2108622 in the model, ancestry explained ∼2.5% of ΔvitE variability; with rs2108622 in the model, ancestry explained ∼1.7% of ΔvitE variability). A consistent direction of association was observed in models of free-cholesterol-adjusted Δα-TOH, but findings were not statistically significant (P = 0.33). There was no evidence for rs2108622 × smoking interaction (P = 0.65). The fully adjusted model explained 18.3% of the variability in plasma Δα-TOH in the vitE arm (*n* = 555): baseline concentrations of α-TOH, γ-TOH, free cholesterol, ancestry, and rs2108622-T explained 9.2%, 1.7%, 1.3%, 1.7%, and 1.3% of the variability, respectively.

## Discussion

We hypothesized that the effect of vitE supplementation on pulmonary function decline would vary across participants in an RCT because of inter-individual variability in the response to vitE supplementation. To address this question, we estimated the longitudinal association of change in vitE status with change in pulmonary function using a dose-response analysis that leveraged a biomarker (change in plasma-measured vitE level) to quantify the treatment exposure. We hypothesized that the associations would be stronger using this more direct measure of treatment exposure compared to the intention-to-treat analysis based on the treatment assignment. Our findings supported the hypothesis, showing that an increase in ΔvitE was associated with an attenuated decline in FEV_1_ in this RCT of generally healthy males ≥50 years of age, particularly in sensitivity analyses focused on adherent vitE supplement responders, such that a 1 SD greater increase in plasma free-cholesterol-adjusted ΔvitE had an effect on FEV_1_ decline that was about one-quarter of the effect of a year of aging, but in the opposite direction. Smoking modified the association such that the beneficial effect of vitE supplementation was statistically significant in never smokers, with less significant effects in current smokers, and little to no effect in former smokers.

To the best of our knowledge, the only prior longitudinal study, which investigated the association of serum α-TOH at study baseline with 8-year decline in pulmonary function, reported that heavy smokers with low baseline serum α-TOH concentration predicted the steepest FEV_1_ decline (24). The previous intention-to-treat analysis in the RAS (25) estimated the treatment effect by study arm and reported a beneficial, but not statistically significant (*P* = 0.19), effect of vitE supplementation on longitudinal pulmonary function; the mean rate of decline in FEV_1_ in the vitE arm was -33 mL/year compared to -39 mL/year in the placebo arm. The early termination of SELECT led to a loss of power in the RAS to detect the intervention effects. The dose-response findings reported herein using the ΔvitE biomarker support a beneficial effect of vitE supplementation on preservation of pulmonary function. These findings warrant further study to consolidate the causal inference and indicate that exploring inter-individual response to nutrition interventions is informative.

Prior studies reported that current smokers experienced a faster disappearance of α-TOH in the circulation and used more α-TOH to combat oxidative stress compared to nonsmokers (47, 48). However, even though we found a significantly lower vitE concentration at baseline in current smokers compared to never and former smokers (19.6 µmol/L, 22.4 µmol/L, and 22.5 µmol/L, respectively, *P* < 0.0005), there was no difference in ΔvitE by smoking status after 3-year vitE supplementation (7.8 µmol/L, 8.0 µmol/L, and 7.7 µmol/L, respectively, *P* > 0.75). Our test of effect modification by smoking status led to the unexpected finding that an increase in vitE attenuated pulmonary function decline in never smokers and current smokers, but not in former smokers. Future studies are needed to replicate the smoking-modified effect observed in this study and, if confirmed, to investigate tissue-specific α-TOH concentrations after vitE supplementation across all smoking groups so as to better understand the mechanism.

We further hypothesized that genetic variation may contribute to the inter-individual variability in ΔvitE. We found that the minor allele (*T*) of rs2108622 on *CYP4F2* was associated with greater ΔvitE (*P* = 0.0032), and this finding partly explained the association of ancestry with ΔvitE. In the fully adjusted model, rs2108622 explained variability in Δα-TOH equivalent to that explained by plasma free cholesterol, which is a key predictor of plasma α-TOH concentration. This *CYP4F2* SNP (rs2108622) passed genome-wide significance in a previous GWAS of ΔvitE conducted in a male heavy smoker population, but the finding was not replicated prior to the study reported herein. Given our population of healthy males, the replication provides important evidence in support of the generalizability of this finding.

The enzyme encoded by the *CYP4F2* gene has been shown to be the only one among the cytochrome P450 enzymes that has *ω*-hydroxylase activity for vitE catabolism (e.g., α- and γ-TOH) in the liver (49, 50). The rs2108622-*T* allele results in a methionine, instead of a valine, at position 433 of the CYP4F2 enzyme, and a prior study showed that the CYP4F2 enzyme goes through faster degradation by the proteasome (51) when methionine is present. If true, persons with the *T* allele would have more α-TOH in the liver available for α-TTP transport into the circulation, leading to higher plasma α-TOH. Consistent with this proposed mechanism, we observed that the EA population, with a higher frequency of rs2108622-*T* minor allele (31% *versus* 13% in AA, similar to those in the 1000 Genomes Project European [29%] and African [8%] populations), had a greater Δα-TOH after supplementation. A candidate gene study of 2 SNPs on the *CYP4F2* gene also reported an association of rs2108622-T with a greater increase in plasma vitE in patients with nonalcoholic fatty liver disease (52).

Several limitations deserve mention. First, at the time of this study, genotype data were available only for participants in the vitE arm (*n* = 555), which limited power to detect significant associations at the genome-wide threshold (*P* < 5×10^−8^). Nevertheless, we replicated one missense SNP (rs2108622) reported in a previous GWAS, surpassing the Bonferroni-corrected p-value threshold for testing two candidate SNPs (*P* < 0.025) (35). Secondly, while our study did not measure all non-genetic factors identified in the prior literature related to plasma vitE, such as waist circumference (WC) and waist-to-hip ratio (WHR) (53), the factors we investigated were representative of the unmeasured ones (e.g., *r* > 0.60 for the correlations of BMI with WC and WHR (54, 55)). Third, the randomized treatment group variable was retained in models because plasma ΔvitE was used to quantify the biological dose of the treatment exposure in the vitE intervention arms rather than the initial intent-to-treat assignment. We found that the association of ΔvitE with pulmonary function was largely unchanged with and without adjusting for potential confounding factors (e.g., age, smoking status, baseline nutriture), although we cannot fully rule out residual confounding. Finally, because the RAS participants were overall healthy males over 50 years old, the generalizability of the study may be limited, and whether the same findings would apply to females, to COPD patients, or to other ancestries beyond EAs and AAs needs further investigation.

In addition, there are several important considerations. First, regarding the timing of the study intervention and the measurement of PFTs, since the RAS was a post-randomization ancillary study to SELECT, no PFT data were available for the pre-intervention time period. Study supplementation was discontinued in October 2008 due to no benefit of the intervention on prostate cancer (36). Although PFTs were measured beyond the end of supplementation, only PFTs measured prior to March 1, 2009 were included in order to provide a conservative estimate of the effect of vitE intervention (4 months beyond the termination of supplements). Moreover, due to the discontinuation of SELECT, 65% of RAS participants completed ≥2 PFTs, leading to a loss of power in the intention-to-treat analysis by not having longitudinal PFT measures available on the full cohort (25). Given that the termination of study supplementation was an independent event unrelated to the timing of annual study visits for the participants, we assumed that the RAS men with ≥2 PFTs comprised a random sample of the entire RAS population. In support of this assumption, the distribution of baseline characteristics including age, height, body mass index (BMI), ancestry, and smoking status were similar comparing men with and without ≥2 PFTs (data not shown). Finally, the findings were consistent in analyses of ΔvitE or cholesterol-adjusted ΔvitE although not all findings were statistically significant.

This study examines both non-genetic and genetic factors related to ΔvitE after vitE supplementation in a generally healthy population, with valid and reproducible measurements of longitudinal plasma vitE status (baseline and year 3, **Supplemental methods**) and pulmonary function (1-5 times per individual). Previous studies investigating the determinants of plasma α-TOH were mostly cross-sectional (53, 56-65) or based on interventions with small numbers of participants (n < 50) (66, 67). The present study is unique in considering ΔvitE in a relatively large sample size (*n* = 1,144), which is important given inter-individual variability in ΔvitE.

To summarize, a greater change in plasma ΔvitE with vitE supplementation was associated with an attenuation in FEV_1_ decline, which may indicate a feasible intervention to mitigate pulmonary function decline. In addition, we replicated one missense variant, rs2108622 in the *CYP4F2* gene, from a previous GWAS of ΔvitE in heavy smokers, which could partially explain variability in ΔvitE after supplementation, as observed in our population of healthy males over 50 years old. This genetic finding contributes to the evidence base for precision nutrition by supporting that genetic subgroups respond differentially to dietary supplementation. Thus, participants with the rs2108622-T minor allele may need less vitE supplement, compared with individuals without this allele, to achieve the same amount of increase in plasma ΔvitE.

## Supporting information

Reporting checklist

## Data Availability

Data described in the manuscript, code book, and analytic code will be made available upon request.

## Acknowledgments

We thank Alison Munson, John Le Barre, and Rachel Grant who were undergraduate research assistants contributing to the analytic work on the association of plasma vitE with pulmonary function and the vitE assay work. We also thank Professor Andrew Clark in the Department of Biological Statistics and Computational Biology at Cornell University for providing advice on the analysis and interpretation of the genetic findings. We would also like to thank Vicky A. Simon in the Human Nutritional Chemistry Service Laboratory at Cornell University for her assistance with the inventory and transferring of the DNA samples. We thank Jing Wu and the Biotechnology Resource Center (BRC) Genomics Facility at Cornell’s Institute of Biotechnology for the quantification and re-adjustment of the DNA sample concentrations. We also want to thank the BRC Bioinformatics Facility for the storage and computational support of the genomic data. In addition, we want to thank Bonnie K. Patchen, PhD candidate in our research group, for providing insight into the interpretation of results. Last but not least, the authors also thank the SELECT/RAS staff, other investigators and the participants of the RAS study for their valuable contributions.

## Authorship’s contribution

JX, KAG, and PAC conceived and designed the study; NCG imputed the genome-wide data; JX, KAG, and NCG analyzed the data. JX, KAG, JMF, ARK, KBA, PJG, CMT, DBH, and PAC interpreted the results; AHA and RSP conducted the vitE assays; PAC, ARK, and CMT designed RAS; PAC, KAG, AHA, and PJG collected data; JX, KAG, DBH, and PAC drafted the manuscript and had primary responsibility for final content; and all authors contributed to critical revision in preparation for publication.

## Data sharing

Data described in the manuscript, code book, and analytic code will be made available upon request.

## Clinical Trial Registry Number

The ClinicalTrials.gov identifier of the SELECT trial is NCT00006392 and the identifier of the RAS trial is NCT00063453.

## References

1. Mocchegiani E, Costarelli L, Giacconi R, Malavolta M, Basso A, Piacenza F, Ostan R, Cevenini E, Gonos ES, Franceschi C, et al. Vitamin E-gene interactions in aging and inflammatory age-related diseases: implications for treatment. A systematic review. Ageing Res Rev. 2014;14:81–101.

2. Brigham KL. Role of free radicals in lung injury. Chest. 1986;89(6):859–63.

3. Dow L, Tracey M, Villar A, Coggon D, Margetts BM, Campbell MJ, Holgate ST. Does dietary intake of vitamins C and E influence lung function in older peopleã Am J Respir Crit Care Med. 1996;154(5):1401–4.

4. Butland BK, Fehily AM, Elwood PC. Diet, lung function, and lung function decline in a cohort of 2512 middle aged men. Thorax. 2000;55(2):102–8.

5. Schunemann HJ, McCann S, Grant BJ, Trevisan M, Muti P, Freudenheim JL. Lung function in relation to intake of carotenoids and other antioxidant vitamins in a population-based study. Am J Epidemiol. 2002;155(5):463–71.

6. Hanson C, Lyden E, Furtado J, Campos H, Sparrow D, Vokonas P, Litonjua AA. Serum tocopherol levels and vitamin E intake are associated with lung function in the normative aging study. Clin Nutr. 2016;35(1):169–74.

7. Joshi P, Kim WJ, Lee SA. The effect of dietary antioxidant on the COPD risk: the community-based KoGES (Ansan-Anseong) cohort. Int J Chron Obstruct Pulmon Dis. 2015;10:2159–68.

8. Charles LE, Burchfiel CM, Mnatsakanova A, Fekedulegn D, Tinney-Zara C, Joseph PN, Schunemann HJ, Violanti JM, Andrew ME, Ochs-Balcom HM. Antioxidants and pulmonary function among police officers. J Occup Environ Med. 2010;52(11):1124–31.

9. Gilliland FD, Berhane KT, Li YF, Gauderman WJ, McConnell R, Peters J. Children’s lung function and antioxidant vitamin, fruit, juice, and vegetable intake. Am J Epidemiol. 2003;158(6):576–84.

10. Tabak C, Smit HA, Rasanen L, Fidanza F, Menotti A, Nissinen A, Feskens EJ, Heederik D, Kromhout D. Dietary factors and pulmonary function: a cross sectional study in middle aged men from three European countries. Thorax. 1999;54(11):1021–6.

11. Hu G, Cassano PA. Antioxidant nutrients and pulmonary function: the Third National Health and Nutrition Examination Survey (NHANES III). Am J Epidemiol. 2000;151(10):975–81.

12. Marchese ME, Kumar R, Colangelo LA, Avila PC, Jacobs DR, Jr., Gross M, Sood A, Liu K, Cook-Mills JM. The vitamin E isoforms alpha-tocopherol and gamma-tocopherol have opposite associations with spirometric parameters: the CARDIA study. Respir Res. 2014;15:31.

13. Schunemann HJ, Grant BJ, Freudenheim JL, Muti P, Browne RW, Drake JA, Klocke RA, Trevisan M. The relation of serum levels of antioxidant vitamins C and E, retinol and carotenoids with pulmonary function in the general population. Am J Respir Crit Care Med. 2001;163(5):1246–55.

14. Grant BJ, Kudalkar DP, Muti P, McCann SE, Trevisan M, Freudenheim JL, Schunemann HJ. Relation between lung function and RBC distribution width in a population-based study. Chest. 2003;124(2):494–500.

15. McKeever TM, Lewis SA, Smit HA, Burney P, Cassano PA, Britton J. A multivariate analysis of serum nutrient levels and lung function. Respir Res. 2008;9:67.

16. Britton JR, Pavord ID, Richards KA, Knox AJ, Wisniewski AF, Lewis SA, Tattersfield AE, Weiss ST. Dietary antioxidant vitamin intake and lung function in the general population. Am J Respir Crit Care Med. 1995;151(5):1383–7.

17. Grievink L, Smit HA, Ocke MC, van ‘t Veer P, Kromhout D. Dietary intake of antioxidant (pro)-vitamins, respiratory symptoms and pulmonary function: the MORGEN study. Thorax. 1998;53(3):166–71.

18. Chen R, Tunstall-Pedoe H, Bolton-Smith C, Hannah MK, Morrison C. Association of dietary antioxidants and waist circumference with pulmonary function and airway obstruction. Am J Epidemiol. 2001;153(2):157–63.

19. Grievink L, de Waart FG, Schouten EG, Kok FJ. Serum carotenoids, alpha-tocopherol, and lung function among Dutch elderly. Am J Respir Crit Care Med. 2000;161(3 Pt 1):790-5.

20. Grievink L, Smit HA, Veer P, Brunekreef B, Kromhout D. Plasma concentrations of the antioxidants beta-carotene and alpha-tocopherol in relation to lung function. Eur J Clin Nutr. 1999;53(10):813–7.

21. Kelly Y, Sacker A, Marmot M. Nutrition and respiratory health in adults: findings from the health survey for Scotland. The European respiratory journal. 2003;21(4):664–71.

22. Molinie F, Favier A, Kauffmann F, Berr C. Effects of lipid peroxidation and antioxidant status on peak flow in a population aged 59-71 years: the EVA study. Respir Med. 2003;97(8):939–46.

23. Pearson P, Britton J, McKeever T, Lewis SA, Weiss S, Pavord I, Fogarty A. Lung function and blood levels of copper, selenium, vitamin C and vitamin E in the general population. Eur J Clin Nutr. 2005;59(9):1043–8.

24. Guenegou A, Leynaert B, Pin I, Le Moel G, Zureik M, Neukirch F. Serum carotenoids, vitamins A and E, and 8 year lung function decline in a general population. Thorax. 2006;61(4):320–6.

25. Cassano PA, Guertin KA, Kristal AR, Ritchie KE, Bertoia ML, Arnold KB, Crowley JJ, Hartline J, Goodman PJ, Tangen CM, et al. A randomized controlled trial of vitamin E and selenium on rate of decline in lung function. Respir Res. 2015;16:35.

26. Klein EA, Thompson IM, Jr., Tangen CM, Crowley JJ, Lucia MS, Goodman PJ, Minasian LM, Ford LG, Parnes HL, Gaziano JM, et al. Vitamin E and the risk of prostate cancer: the Selenium and Vitamin E Cancer Prevention Trial (SELECT). Jama. 2011;306(14):1549–56.

27. Alpha-Tocopherol BCCPSG. The effect of vitamin E and beta carotene on the incidence of lung cancer and other cancers in male smokers. N Engl J Med. 1994;330(15):1029–35.

28. Heart Outcomes Prevention Evaluation Study I, Yusuf S, Dagenais G, Pogue J, Bosch J, Sleight P. Vitamin E supplementation and cardiovascular events in high-risk patients. N Engl J Med. 2000;342(3):154–60.

29. Gupta SK. Intention-to-treat concept: A review. Perspect Clin Res. 2011;2(3):109–12.

30. Kussmann M, Fay LB. Nutrigenomics and personalized nutrition: science and concept. Per Med. 2008;5(5):447–55.

31. Doring F, Rimbach G, Lodge JK. In silico search for single nucleotide polymorphisms in genes important in vitamin E homeostasis. IUBMB Life. 2004;56(10):615–20.

32. Ferrucci L, Perry JR, Matteini A, Perola M, Tanaka T, Silander K, Rice N, Melzer D, Murray A, Cluett C, et al. Common variation in the beta-carotene 15,15’-monooxygenase 1 gene affects circulating levels of carotenoids: a genome-wide association study. Am J Hum Genet. 2009;84(2):123–33.

33. Major JM, Yu K, Wheeler W, Zhang H, Cornelis MC, Wright ME, Yeager M, Snyder K, Weinstein SJ, Mondul A, et al. Genome-wide association study identifies common variants associated with circulating vitamin E levels. Hum Mol Genet. 2011;20(19):3876–83.

34. Wood AR, Perry JR, Tanaka T, Hernandez DG, Zheng HF, Melzer D, Gibbs JR, Nalls MA, Weedon MN, Spector TD, et al. Imputation of variants from the 1000 Genomes Project modestly improves known associations and can identify low-frequency variant-phenotype associations undetected by HapMap based imputation. PloS one. 2013;8(5):e64343.

35. Major JM, Yu K, Chung CC, Weinstein SJ, Yeager M, Wheeler W, Snyder K, Wright ME, Virtamo J, Chanock S, et al. Genome-wide association study identifies three common variants associated with serologic response to vitamin E supplementation in men. J Nutr. 2012;142(5):866–71.

36. Lippman SM, Klein EA, Goodman PJ, Lucia MS, Thompson IM, Ford LG, Parnes HL, Minasian LM, Gaziano JM, Hartline JA, et al. Effect of selenium and vitamin E on risk of prostate cancer and other cancers: the Selenium and Vitamin E Cancer Prevention Trial (SELECT). Jama. 2009;301(1):39–51.

37. Barr RG, Stemple KJ, Mesia-Vela S, Basner RC, Derk SJ, Henneberger PK, Milton DK, Taveras B. Reproducibility and validity of a handheld spirometer. Respir Care. 2008;53(4):433–41.

38. Miller MR, Hankinson J, Brusasco V, Burgos F, Casaburi R, Coates A, Crapo R, Enright P, van der Grinten CP, Gustafsson P, et al. Standardisation of spirometry. The European respiratory journal. 2005;26(2):319–38.

39. Lopez A, Krehl WA, Hodges RE. Relationship between total cholesterol and cholesteryl esters with age in human blood plasma. Am J Clin Nutr. 1967;20(8):808–15.

40. Ford ES, Schleicher RL, Mokdad AH, Ajani UA, Liu S. Distribution of serum concentrations of alpha-tocopherol and gamma-tocopherol in the US population. Am J Clin Nutr. 2006;84(2):375–83.

41. Das S, Forer L, Schonherr S, Sidore C, Locke AE, Kwong A, Vrieze SI, Chew EY, Levy S, McGue M, et al. Next-generation genotype imputation service and methods. Nature genetics. 2016;48(10):1284–7.

42. Fred Hutch Food Frequency Questionnaires (FFQ) [Available from: https://sharedresources.fredhutch.org/services/food-frequency-questionnaires-ffq.

43. Marchini J, Howie B. Genotype imputation for genome-wide association studies. Nat Rev Genet. 2010;11(7):499–511.

44. Zhan X, Hu Y, Li B, Abecasis GR, Liu DJ. RVTESTS: an efficient and comprehensive tool for rare variant association analysis using sequence data. Bioinformatics. 2016;32(9):1423–6.

45. Willer CJ, Li Y, Abecasis GR. METAL: fast and efficient meta-analysis of genomewide association scans. Bioinformatics. 2010;26(17):2190–1.

46. Institute of Medicine (US). Panel on Dietary Antioxidants and Related Compounds. Dietary Reference Intakes for Vitamin C, Vitamin E, Selenium, and Carotenoids: A Report of the Panel on Dietary Antioxidants and Related Compounds. Washington, D.C.: National Academy Press; 2000.

47. Traber MG, Winklhofer-Roob BM, Roob JM, Khoschsorur G, Aigner R, Cross C, Ramakrishnan R, Brigelius-Flohe R. Vitamin E kinetics in smokers and nonsmokers. Free Radic Biol Med. 2001;31(11):1368–74.

48. Bruno RS, Ramakrishnan R, Montine TJ, Bray TM, Traber MG. {alpha}-Tocopherol disappearance is faster in cigarette smokers and is inversely related to their ascorbic acid status. Am J Clin Nutr. 2005;81(1):95–103.

49. Sontag TJ, Parker RS. Cytochrome P450 omega-hydroxylase pathway of tocopherol catabolism. Novel mechanism of regulation of vitamin E status. J Biol Chem. 2002;277(28):25290–6.

50. Parker RS, Sontag TJ, Swanson JE, McCormick CC. Discovery, characterization, and significance of the cytochrome P450 omega-hydroxylase pathway of vitamin E catabolism. Ann N Y Acad Sci. 2004;1031:13–21.

51. van Engen CE, Ofman R, Dijkstra IM, van Goethem TJ, Verheij E, Varin J, Vidaud M, Wanders RJ, Aubourg P, Kemp S, et al. CYP4F2 affects phenotypic outcome in adrenoleukodystrophy by modulating the clearance of very long-chain fatty acids. Biochim Biophys Acta. 2016;1862(10):1861–70.

52. Athinarayanan S, Wei R, Zhang M, Bai S, Traber MG, Yates K, Cummings OW, Molleston J, Liu W, Chalasani N. Genetic polymorphism of cytochrome P450 4F2, vitamin E level and histological response in adults and children with nonalcoholic fatty liver disease who participated in PIVENS and TONIC clinical trials. PloS one. 2014;9(4):e95366.

53. Wallstrom P, Wirfalt E, Lahmann PH, Gullberg B, Janzon L, Berglund G. Serum concentrations of beta-carotene and alpha-tocopherol are associated with diet, smoking, and general and central adiposity. Am J Clin Nutr. 2001;73(4):777–85.

54. Vazquez G, Duval S, Jacobs DR, Jr., Silventoinen K. Comparison of body mass index, waist circumference, and waist/hip ratio in predicting incident diabetes: a meta-analysis. Epidemiol Rev. 2007;29:115–28.

55. Ohlson LO, Larsson B, Svardsudd K, Welin L, Eriksson H, Wilhelmsen L, Bjorntorp P, Tibblin G. The influence of body fat distribution on the incidence of diabetes mellitus. 13.5 years of follow-up of the participants in the study of men born in 1913. Diabetes. 1985;34(10):1055–8.

56. Abdulla KA, Um CY, Gross MD, Bostick RM. Circulating gamma-Tocopherol Concentrations Are Inversely Associated with Antioxidant Exposures and Directly Associated with Systemic Oxidative Stress and Inflammation in Adults. J Nutr. 2018;148(9):1453–61.

57. Waniek S, di Giuseppe R, Esatbeyoglu T, Plachta-Danielzik S, Ratjen I, Jacobs G, Nothlings U, Koch M, Schlesinger S, Rimbach G, et al. Vitamin E (alpha- and gamma-Tocopherol) Levels in the Community: Distribution, Clinical and Biochemical Correlates, and Association with Dietary Patterns. Nutrients. 2017;10(1).

58. Stuetz W, Weber D, Dolle ME, Jansen E, Grubeck-Loebenstein B, Fiegl S, Toussaint O, Bernhardt J, Gonos ES, Franceschi C, et al. Plasma Carotenoids, Tocopherols, and Retinol in the Age-Stratified (35-74 Years) General Population: A Cross-Sectional Study in Six European Countries. Nutrients. 2016;8(10).

59. Talegawkar SA, Johnson EJ, Carithers T, Taylor HA, Jr., Bogle ML, Tucker KL. Total alpha-tocopherol intakes are associated with serum alpha-tocopherol concentrations in African American adults. J Nutr. 2007;137(10):2297–303.

60. Galan P, Viteri FE, Bertrais S, Czernichow S, Faure H, Arnaud J, Ruffieux D, Chenal S, Arnault N, Favier A, et al. Serum concentrations of beta-carotene, vitamins C and E, zinc and selenium are influenced by sex, age, diet, smoking status, alcohol consumption and corpulence in a general French adult population. Eur J Clin Nutr. 2005;59(10):1181–90.

61. White E, Kristal AR, Shikany JM, Wilson AC, Chen C, Mares-Perlman JA, Masaki KH, Caan BJ. Correlates of serum alpha- and gamma-tocopherol in the Women’s Health Initiative. Ann Epidemiol. 2001;11(2):136–44.

62. Vogel S, Contois JH, Tucker KL, Wilson PW, Schaefer EJ, Lammi-Keefe CJ. Plasma retinol and plasma and lipoprotein tocopherol and carotenoid concentrations in healthy elderly participants of the Framingham Heart Study. Am J Clin Nutr. 1997;66(4):950–8.

63. Ascherio A, Stampfer MJ, Colditz GA, Rimm EB, Litin L, Willett WC. Correlations of vitamin A and E intakes with the plasma concentrations of carotenoids and tocopherols among American men and women. J Nutr. 1992;122(9):1792–801.

64. Knekt P, Seppanen R, Aaran RK. Determinants of serum alpha-tocopherol in Finnish adults. Prev Med. 1988;17(6):725–35.

65. Willett WC, Stampfer MJ, Underwood BA, Speizer FE, Rosner B, Hennekens CH. Validation of a dietary questionnaire with plasma carotenoid and alpha-tocopherol levels. Am J Clin Nutr. 1983;38(4):631–9.

66. Switzer BR, Atwood JR, Stark AH, Hatch JW, Travis R, Ullrich F, Lyden ER, Wu X, Chiu Y, Smith JL. Plasma carotenoid and vitamins a and e concentrations in older African American women after wheat bran supplementation: effects of age, body mass and smoking history. J Am Coll Nutr. 2005;24(3):217–26.

67. Grolier P, Boirie Y, Levadoux E, Brandolini M, Borel P, Azais-Braesco V, Beaufrere B, Ritz P. Age-related changes in plasma lycopene concentrations, but not in vitamin E, are associated with fat mass. Br J Nutr. 2000;84(5):711–6.

