## Supplemental Material for "Change in Plasma Alpha-Tocopherol Associations with Attenuated Pulmonary Function Decline and with *CYP4F2* Missense Variation"

***Supplemental Methods***

*RAS Participants*

RAS enrolled 2,921 participants, and 2,846 participants had $\geq$ 1 pulmonary function measurement that met American Thoracic Society (ATS) standardization of spirometry guidelines (1). Among RAS participants who had pulmonary function measured (*n* = 2,846), 74 participants were lost to follow-up (~2.6%).

*Vitamin E and Free Cholesterol Measurements*

Samples were assayed by the same technician blinded to the sample identity and study arm, with a control sample assayed in the first and last run every day; assays were completed over a 6-month period beginning in June 2010. Both the tocopherols (TOHs) and free cholesterol (as silyl ethers) concentrations were quantitated against the internal standard of d9-α-TOH, with cholesterol adjusted for differences in detector responses. All samples were assayed in batches of eight samples (four paired samples for each individual at baseline and year 3) with one duplicate sample for one individual per batch, which generated a total of 321 duplicated sets. For all the plasma samples, the mean within-batch coefficient of variation (CV) was 9.6% for total α-TOH and 1.7% for free cholesterol-adjusted α-TOH, whereas the CV for γ-TOH was 13.4%, which suggested a greater variation in γ-TOH relative to its mean, compared to α-TOH. For the control samples, the mean between-run CV was 13.9% for α-TOH and 12.9% for cholesterol-adjusted α-TOH with standard deviation of 6.5 μmol/L and 2.1 μmol/mmol free cholesterol for α-TOH and free-cholesterol-adjusted α-TOH, respectively.

When measuring the free cholesterol concentrations, there was a loss of sensitivity of the gas chromatography column, which was addressed by rerunning and applying a correction factor to all affected samples. Given the consistent findings observed across the 2 ΔvitE phenotypes (raw or free-cholesterol-adjusted), the impact of this issue is expected to be minor.

The average α-TOH concentration at baseline in the 2 vitamin E arms in RAS was 21.9 µmol/L, which was slightly lower, but fairly close to the average α-TOH concentration (26.2 µmol/L) among the male population in the United States based on the third National Health and Nutrition Examination Survey (2). In addition, the slightly smaller mean could possibly be due to the fact that participants included in RAS had stopped taking vitamin E supplementation when they were enrolled at baseline whereas the NHANES population included individuals who took vitamin E supplementation at the time of the survey (3). Thus, the likelihood of degradation of the vitamin E assay with a maximum of 9-year storage time should be minimal.

*Non-Genetic Factors in Relation to Plasma Response to Supplementation*

Previous literature was searched to identify biologically plausible associations of other nutrients with vitE. Plasma selenium concentration at baseline was included given its role in reducing lipid hydroperoxides and its potential synergistic effect with vitE through the glutathione-peroxidase system (4, 5). Vitamin C (vitC) can regenerate vitE from the reaction with lipid hydroperoxyl radicals (6), however, our study did not measure plasma vitC concentration. We adjusted for dietary intake of vitC as a proxy for vitC status in this study; a prior meta-analysis reported a moderate correlation between dietary and plasma vitC (*r* = 0.46) in males (7), and thus provided a rationale for this approach. Alcohol was also included as a potential factor that might affect plasma vitE status, given the antioxidant effect of vitE to alleviate the oxidative stress induced by alcohol in the liver (8). Dietary fiber was considered given the mechanistic evidence that fiber binds to vitE in the intestine and therefore vitE absorption may be reduced in the presence of a diet higher in fiber (9, 10).

**Supplemental Table 1. Plasma tocopherol concentrations in male participants in the two vitamin E arms as well as in the placebo arm of the Respiratory Ancillary Study ^1^**

|  | Baseline | | | | Change after 3-year supplementation ^2^ | | | |
| --- | --- | --- | --- | --- | --- | --- | --- | --- |
|  | Double Placebo  (*n* = 91) | Vitamin E + Selenium  (*n* = 565) | Vitamin E + Placebo  (*n* = 579) | P_vitE+Se_  _vs vitE_ ^3^ | Double Placebo  (*n* = 91) | Vitamin E + Selenium  (*n* = 565) | Vitamin E + Placebo  (*n* = 579) | P_vitE+Se_  _vs vitE_ ^3^ |
| α-tocopherol |  |  |  |  |  |  |  |  |
| Unadjusted (µmol/L) | 23.1 (11.5) | 22.3 (9.8) | 21.5 (9.0) | 0.16 | -3.8 (8.5) | 7.4 (14.5) | 8.2 (12.5) | 0.28 |
| Free-cholesterol-adjusted  (µmol/mmol chol) | 9.9 (3.1) | 9.3 (3.5) | 9.1 (3.1) | 0.38 | -1.1 (2.4) | 3.6 (4.7) | 3.5 (4.1) | 0.77 |
| γ-tocopherol |  |  |  |  |  |  |  |  |
| Unadjusted (µmol/L) | 3.3 (2.9) | 3.4 (2.6) | 3.5 (2.4) | 0.43 | 0.5 (2.9) | -1.7 (2.6) | -1.7 (2.4) | 0.63 |
| Free-cholesterol-adjusted  (µmol/mmol chol) | 1.4 (0.9) | 1.4 (0.9) | 1.5 (0.9) | 0.054 | 0.2 (0.9) | -0.7 (0.9) | -0.7 (0.9) | 0.60 |
| Vitamin E deficiency,  No. (%) ^4^ | 6 (6.6%) | 31 (5.5%) | 37 (6.4%) | 0.52 | 11 (12.1%) | 9 (1.6%) | 13 (2.2%) | 0.42 |

^1^ Data are presented as mean (standard deviation [SD]) in the table, unless otherwise noted. Vitamin E supplementation refers to 400 IU/day *all rac-α*-tocopherol, and includes participants randomized to vitamin E + selenium and to vitamin E + placebo.

^2^ Change in tocopherol concentration was calculated as tocopherol concentration at baseline (or pre-supplementation) subtracted from concentration at year 3. Change in free-cholesterol-adjusted tocopherol concentration was calculated as the difference between the tocopherol concentration at year 3 divided by free cholesterol concentration at year 3 and the tocopherol concentration at baseline divided by free cholesterol concentration at baseline.

^3^ P-value for the test of the difference in mean tocopherol (or frequency of vitamin E deficiency) in the vitE+Se arm versus the vitE arm.

^4^ At risk of vitamin E deficiency defined as <12 μmol/L α-tocopherol (11) at study baseline and at year 3.

Abbreviations: µmol/mmol chol, µmol/mmol cholesterol; vitE, vitamin E supplementation only arm; vitE+Se, vitamin E + selenium supplementation arm.

**Supplemental Table 2. Association of plasma change in γ-tocopherol after 3-year vitamin E supplementation with annual rate of change in FEV_1_ ^1^**

|  | $\boldsymbol{\Delta}$Raw γ-TOH | | | $\boldsymbol{\Delta}$Adjusted γ-TOH ^2^ | | |
| --- | --- | --- | --- | --- | --- | --- |
|  | $\beta$ (SE) ^3^ | 95% CI | P value ^4^ | $\beta$ (SE) ^3^ | 95% CI | P value ^4^ |
| Main effect model |  |  |  |  |  |  |
| - Full sample (*n* = 1,142^1^) | -0.52 (1.06) | (-2.61, 1.56) | 0.62 | -5.46 (2.97) | (-11.29, 0.37) | 0.067 |
| - Adherent sample ^5^ (*n* = 775) | -0.33 (1.32) | (-2.91, 2.25) | 0.80 | -5.68 (3.76) | (-13.07, 1.72) | 0.13 |
| - Adherent sample who responded to the vitE supplement ^5,6^ (*n* = 680) | 0.16 (1.43) | (-2.65, 2.98) | 0.91 | -6.22 (4.11) | (-14.29, 1.85) | 0.13 |

^1^ All models were adjusted for age at baseline, height, ancestry, smoking status, smoking dose at baseline, treatment arm, baseline α-tocopherol, baseline γ-tocopherol, and baseline free cholesterol (only in the model with raw γ-tocopherol values). 2 participants had missing data for smoking dose and thus the sample size for the fully adjusted model was 1,142 out of 1,144 participants in the vitamin E + selenium arm and in the vitamin E + placebo arm.

^2^ $\Delta$Adjusted γ-TOH refers to the change in free-cholesterol-adjusted γ-tocopherol concentration from baseline to year 3.

^3^ $\beta$ is the coefficient for the $\Delta$ γ-TOH $\times$ time term in the main effect model.

^4^ P-value of the $\Delta$ γ-TOH $\times$ time term in the model adjusted for other covariates, which indicates whether the coefficient significantly differed from 0. Significant *P <* 0.05 is in bold.

^5^ Adherent participants (defined as taking at least 80% of the supplement pills) during the 3 years of supplementation.

^6^ Responder was defined as $\Delta$raw α-TOH greater than the mean change in the placebo arm (-3.8 µmol/L) after 3-year supplementation.

Abbreviations: CI, confidence interval.

**Supplemental Table 3. Association of the interaction of plasma change in α-tocopherol after 3-year vitamin E supplementation with smoking status on annual rate of change in FEV_1_ in adherent participants in the vitamin E arms (*n* = 775) ^1^**

|  | $\boldsymbol{\Delta}$Raw α-TOH | | $\boldsymbol{\Delta}$Adjusted α-TOH ^2^ | |
| --- | --- | --- | --- | --- |
|  | $\beta$ (SE) ^3^ | P ^4^ | $\beta$ (SE) ^3^ | P ^4^ |
| Smoking interaction model |  | P_interaction_=0.082 ^5^ |  | P_interaction_=0.12 ^5^ |
| - Never smokers (*n* = 242) | 0.71 (0.43) | 0.095 | 2.90 (1.30) | **0.026** |
| - Former smokers (*n* = 420) | -0.33 (0.36) | 0.35 | -0.29 (1.11) | 0.79 |
| - Current smokers (*n* = 113) | 0.97 (0.65) | 0.14 | 3.02 (2.04) | 0.14 |

^1^ All adherent participants (defined as taking at least 80% of the supplement pills) in the vitamin E + selenium arm and in the vitamin E + placebo arm. The model has been adjusted for age at baseline, height, ancestry, smoking status, smoking dose at baseline, treatment arm, baseline α-tocopherol, baseline γ-tocopherol, and baseline free cholesterol (only in the model with raw α-tocopherol).

^2^ $\Delta$Adjusted α-TOH refers to the change in free-cholesterol-adjusted α-tocopherol concentration from baseline to year 3.

^3^ $\beta$ is the coefficient of the $\Delta$α-TOH $\times$ time term in the statistical model for each smoking stratum.

^4^ P-value of the $\Delta$α-TOH $\times$ time term for each smoking group (when it was set as the reference) in the model that included all smoking status. Significant *P <* 0.05 is in bold.

^5^ The P_interaction_ value was for the 3-way interaction term (∆α-TOH × time × smoking status) to test whether the association of $\Delta$α-TOH on rate of change in FEV_1_ differed by smoking status.

**Supplemental Table 4. Plasma change in α-tocopherol by cigarette smoking status in the two vitamin E arms in the Respiratory Ancillary Study (*n* = 1,144) ^1^**

|  | $\boldsymbol{\Delta}$Raw α-TOH  (µmol/L) | | | | | |
| --- | --- | --- | --- | --- | --- | --- |
|  | EA participants  in the vitE arm  (*n* = 439) | AA participants  in the vitE arm  (*n* = 140) | P_EA vs AA_ ^2^ | EA participants  in the vitE+Se arm  (*n* = 435) | AA participants  in the vitE+Se arm  (*n* = 130) | P_EA vs AA_ ^2^ |
| Never smoker | 7.8 (12.9) | 6.5 (11.2) | 0.58 | 7.7 (14.6) | 11.6 (12.0) | 0.17 |
| Former smoker | 9.5 (12.3) | 4.7 (13.7) | **0.0087** | 7.3 (15.9) | 5.3 (10.3) | 0.28 |
| Current smoker | 10.1 (12.5) | 7.0 (10.6) | 0.17 | 8.0 (13.5) | 5.4 (12.9) | 0.31 |

^1^ Data are presented as mean (standard deviation) in the table, unless otherwise noted. Change in tocopherol concentration was calculated as tocopherol concentration at baseline (or pre-supplementation) subtracted from that at year 3; positive values represent increases in tocopherol from baseline to year three.

^2^ P value is the significance level for difference in mean plasma change in α-TOH across EA and AA participants. Significant *P <* 0.05 is in bold.

Abbreviations: AA, African ancestry; EA, European ancestry; vitE, vitamin E supplementation only arm; vitE+Se, vitamin E + selenium supplementation arm.

***Supplemental References***

1.	Miller MR, Hankinson J, Brusasco V, Burgos F, Casaburi R, Coates A, Crapo R, Enright P, van der Grinten CP, Gustafsson P, et al. Standardisation of spirometry. The European respiratory journal. 2005;26(2):319-38.

2.	Ford ES, Sowell A. Serum alpha-tocopherol status in the United States population: findings from the Third National Health and Nutrition Examination Survey. Am J Epidemiol. 1999;150(3):290-300.

3.	Radimer K, Bindewald B, Hughes J, Ervin B, Swanson C, Picciano MF. Dietary supplement use by US adults: data from the National Health and Nutrition Examination Survey, 1999-2000. Am J Epidemiol. 2004;160(4):339-49.

4.	Tappel AL. Vitamin E and selenium protection from in vivo lipid peroxidation. Ann N Y Acad Sci. 1980;355:18-31.

5.	Tappel AL. Selenium-glutathione peroxidase and vitamin E. Am J Clin Nutr. 1974;27(9):960-5.

6.	Niki E. Interaction of ascorbate and alpha-tocopherol. Ann N Y Acad Sci. 1987;498:186-99.

7.	Dehghan M, Akhtar-Danesh N, McMillan CR, Thabane L. Is plasma vitamin C an appropriate biomarker of vitamin C intake? A systematic review and meta-analysis. Nutr J. 2007;6:41.

8.	Kaur J, Shalini S, Bansal MP. Influence of vitamin E on alcohol-induced changes in antioxidant defenses in mice liver. Toxicol Mech Methods. 2010;20(2):82-9.

9.	Nnanna IA, O'Neill KL. In vitro binding of vitamin E to selected dietary fiber sources. J Food Sci. 1992;57(3):721-5.

10.	Palafox-Carlos H, Ayala-Zavala JF, Gonzalez-Aguilar GA. The role of dietary fiber in the bioaccessibility and bioavailability of fruit and vegetable antioxidants. J Food Sci. 2011;76(1):R6-R15.

11.	Institute of Medicine (US). Panel on Dietary Antioxidants and Related Compounds. Dietary Reference Intakes for Vitamin C, Vitamin E, Selenium, and Carotenoids: A Report of the Panel on Dietary Antioxidants and Related Compounds. Washington, D.C.: National Academy Press; 2000.
